# PRDM1 DNA-binding zinc finger domain is required for normal limb development and is disrupted in Split Hand/Foot Malformation

**DOI:** 10.1101/2022.11.16.22282191

**Authors:** Brittany T. Truong, Lomeli C. Shull, Ezra Lencer, Eric G. Bend, Michael Field, University of Washington Center for Mendelian Genomics (UW-CMG), David Everman, Charles E. Schwartz, Heather Flanagan-Steet, Kristin B. Artinger

## Abstract

Split Hand/Foot Malformation (SHFM) is a rare limb abnormality with clefting of the fingers and/or toes. For many patients, the genetic etiology is unknown. Through whole exome and targeted sequencing, we detected three novel variants in a transcription factor, *PRDM1* that arose *de novo* in families with SHFM or segregated with the phenotype. PRDM1 is required for limb development; however, its role is not well understood, and it is unclear how the *PRDM1* variants affect protein function. Using transient and stable overexpression rescue experiments in zebrafish, we show that the variants, which disrupt the proline/serine-rich and DNA-binding zinc finger domains have reduced function compared to wildtype *PRDM1*. Through gene expression assays, RNA-seq, and CUT&RUN in isolated pectoral fin cells, we demonstrate that Prdm1a directly binds to and regulates genes required for limb induction, outgrowth, and anterior/posterior patterning, such as *fgfr1a, dlx5a, dlx6a*, and *smo*. Together, these results improve our understanding of the role of PRDM1 in the limb gene regulatory network and demonstrate the pathogenicity of *PRDM1* variants in humans.

**SUMMARY STATEMENT:** PRDM1 proline/serine and zinc finger domains are required to regulate limb induction, outgrowth, and anterior/posterior patterning. Variants in PRDM1 are shown to cause Split Hand/Foot Malformation in humans.

## INTRODUCTION

Vertebrate limb development is controlled by a complex gene regulatory network (GRN) governed by signaling pathways, transcription factors, and epigenetic modifiers. Limb growth begins at the lateral plate mesoderm, where mesenchyme precursors form a small bud surrounded by an ectodermal layer. Retinoic acid and Wnt signaling initiate limb induction (Grandel et al., 2002; Ng et al., 2002), and outgrowth is driven by the apical ectodermal ridge (AER), where transcription factors TBX5 and TP63 induce expression of fibroblast growth factor 10 (*Fgf10*) in the mesenchyme and *Fgf8* in the outer ectoderm (Agarwal et al., 2003; Bakkers et al., 2002; Ng et al., 2002). This establishes a complex epithelial-mesenchymal feedback loop that then activates proliferation and differentiation of mesenchymal cells for limb growth (proximal/distal axis) (Ohuchi et al., 1997). Anterior/posterior patterning, or establishment of digits 1-5, is regulated by sonic hedgehog (Shh) signaling in the zone of polarizing activity (ZPA) (Riddle et al., 1993; Saunders & Gasseling, 1968). Each gene and pathway are interconnected, and dysregulation at any point, particularly in the AER, can cause abnormal limb growth (Kantaputra & Carlson, 2019).

Misregulation of the limb GRN can lead to congenital limb defects, which affect 1 in 2,000 newborns (Wilcox et al., 2015). Split Hand/Foot Malformation (SHFM) is a limb abnormality resulting in missing, curved, and/or clefted digits. SHFM occurs in 1 in 18,000 live births, and there are six known isotypes of the disease due to pathogenic variants in *WNT10B* (MIM #225300), *TP63* (MIM #605289), and *DLX5* (MIM #225300) or chromosomal rearrangements in chromosomes 2 (MIM %606708), 10 (MIM #246560), and X (MIM %313350); however, in 50% of cases, the genetic etiology is unknown (Sowinska-Seidler et al., 2014). Deletions and translocations at 6q21 have also been associated with SHFM, though no candidate gene has been isolated prior to now (Braverman et al., 1993; Correa-Cerro et al., 1996; Duran-Gonzalez et al., 2007; Gurrieri et al., 1995; Hopkin et al., 1997; Pandya et al., 1995; Tsukahara et al., 1997; Viljoen & Smart, 1993). Here, we report three families with SHFM of unknown genetic etiology, and using whole exome sequencing (WES) and targeted sequencing, we identified three different variants of unknown significance in a transcription factor, *PRDM1*, located at 6q21.

PRDM1, also known as BLIMP1 (MIM *603423), is required for limb development, though its role not well understood (Ha & Riddle, 2003; Lee & Roy, 2006; Mercader et al., 2006; Robertson et al., 2007; Vincent et al., 2005; Wilm & Solnica-Krezel, 2005). The protein has an N-terminal SET domain, followed by a proline/serine-rich domain, and five zinc fingers. In various contexts, PRDM1 can bind to DNA through its zinc finger domains and activate or repress gene expression (reviewed in Bikoff et al. (2009); (Powell et al., 2013)). SET domains are often associated with histone methyltransferase activity (Cheng et al., 2005; Martin & Zhang, 2005), but this has not been observed for PRDM1 *in vivo* (Hohenauer & Moore, 2012). Rather, it can indirectly alter transcription by forming complexes with chromatin-modifying proteins, such as histone demethylase Kdm4a (Prajapati et al., 2019), histone methyltransferases Prmt5 (Ancelin et al., 2006) and G9a (Gyory et al., 2004) and histone deacetylases HDAC1/2 (Yu et al., 2000) at both the pro/ser and zinc finger domains (reviewed in Bikoff et al. (2009)). Studies in mice and zebrafish indicate that PRDM1 is important for limb formation. Conditional knockouts of *Prdm1* in the embryo proper (*Sox2:Cre*) causes loss of posterior digits in mice due to disruption of sonic hedgehog signaling and misregulation in the ZPA (Robertson et al., 2007). Zebrafish embryos injected with *prdm1a* morpholinos “morphants”) for targeted knockdown fail to develop a pectoral fin, which is homologous to mammalian forelimbs, while a hypomorphic allele, *prdm1a*^*tp39/tp39*^, presents with mild phenotypes, namely shortening of the scapulocoracoid and variable truncation of the fin overall (Lee & Roy, 2006; Mercader et al., 2006). These studies show that PRDM1 is downstream of *tbx5a* and upstream of *fgf10a* during fin induction. It is also required for *shh* activity in the ZPA (Lee & Roy, 2006; Mercader et al., 2006). However, it is unclear whether this is by direct transcriptional regulation or by recruitment of epigenetic modifiers. While PRDM1 has been shown to be important in limb development, how it functions molecularly is poorly understood.

In this study, we sought to better understand the mechanistic role of PRDM1 in limb development. We identify novel *PRDM1* variants in SHFM patients and show through transient and stable overexpression assays in zebrafish that the variants have reduced function compared to wildtype PRDM1 due to disruption of the pro/ser and DNA-binding zinc finger domains. We use RNA-seq and CUT&RUN in isolated pectoral fin cells to show that Prdm1a directly binds to *fgfr1a, dlx5a, dlx6a*, and *smo* and regulates their expression in the fin. These data show that PRDM1 is involved in limb induction, outgrowth, and anterior/posterior patterning and requires its pro/ser and zinc finger domains to accomplish these morphogenic processes. Together, these results improve our understanding of the role of PRDM1 in the limb gene regulatory network and demonstrate the pathogenicity of *PRDM1* variants in humans.

## RESULTS

### Whole exome sequencing in SHFM patients reveals novel *PRDM1* variants of unknown significance

SHFM is a congenital limb disorder, in which patients exhibit missing, shortened, or clefting of the fingers and toes. Phenotypes vary in terms of severity as well as bilateral symmetry. For 50% of individuals, the genetic etiology is unknown. We performed WES on a multi-generational family with SHFM that was negative for known SHFM associated genes and exhibited no chromosomal rearrangements cytogenetically or by microarray analyses. Four individuals were heterozygous for a mutation in *PRDM1, PRDM1c*.*712_713insT* (p.C239Lfs*32), which introduces a single base pair insertion causing a frameshift and premature stop codon after the SET domain as well as predicted truncation of the protein (**Fig. 1A-B**). The pLI score (probability of being loss-of-function intolerant) for *PRDM1* on gnomAD is 0.96, suggesting the gene is intolerant to loss-of-function variants. This variant also has a minor allele frequency (MAF) of 0 in gnomAD, is predicted to be pathogenic by MutationTaster (www.mutationtaster.org), and is the only gene from the WES results involved in limb development (**Fig. 1A; Table S1**) (Ha & Riddle, 2003; Lee & Roy, 2006; Mercader et al., 2006; Robertson et al., 2007; Vincent et al., 2005; Wilm & Solnica-Krezel, 2005).

**Figure 1.**
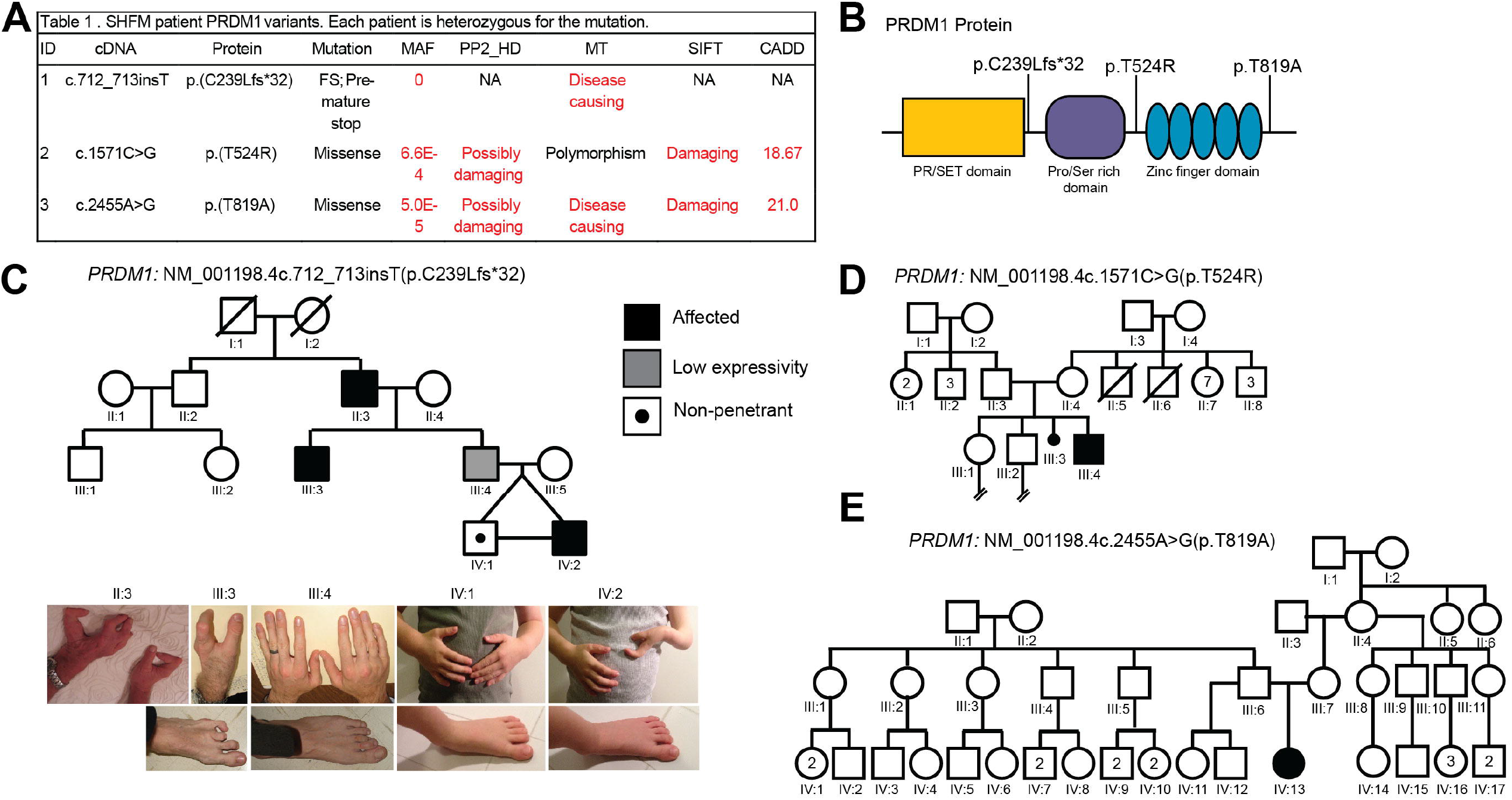
PRDM1 variants of unknown significance identified in families with Split Hand/Foot Malformation (SHFM). (A) Table showing *PRDM1* variants and predictions of pathogenicity based on various bioinformatics tools. (**B**) Schematic of PRDM1 structure and location of SHFM variants identified in human patients. (**C**) Pedigree for family with *PRDM1* variant #1, *c*.*712_713insT* (p.C239Lfs*32). Affected individuals are shaded. Standard pedigree symbols are used. The variant is inherited in an autosomal dominant manner with incomplete penetrance and variable expressivity. Photographs of individuals in the family are also shown. (**D**) Pedigree for family with *PRDM1* variant #2, *c*.*1571C>G* (p.T524R). (**E**) Pedigree for family with *PRDM1* variant #3, *c*.*2455A>G* (p.T819A). Abbreviations: CADD, Combined Annotation-Dependent Depletion; fs, frameshift; HD, HumDiv; maf, minor allele frequency; MT, MutationTaster; PP2, PolyPhen2; SIFT, Sorting Intolerant From Tolerant

Phenotypes for the individuals in this family who carry the variant differ in severity. Individuals II:3 and III:3 have missing and curved fingers but normal feet. Individual III:3 has clefted digits and a missing toe. Individual III:4 has a mild phenotype of minor brachydactyly, or shortened digits. Individuals IV:1 and IV:2 are monozygotic twins, though only IV:2 has SHFM (missing digits and clefting) (**Fig. 1C**). This is likely due to environmental differences while *in utero* and/or epigenetic differences (Castillo-Fernandez et al., 2014; Gordon et al., 2012). This form of SHFM is inherited in an autosomal dominant manner with variable penetrance and expressivity.

We then screened an additional 75 unrelated SHFM patients. Two individuals were negative for any *TP63* variants or chromosomal abnormalities, but we identified two additional missense variants in *PRDM1*. The second variant is *PRDM1c*.*1571C>G* (p.T524R) and the third is *PRDM1c*.*2455A>G* (p.T819A) (**Fig. 1A**). Individuals were heterozygous, and the variants were absent from both sets of parents based on targeted sequencing, suggesting a *de novo* mutation (**Fig. 1D-E**). Both variants have a MAF of ≤6.6E-4 (gnomAD) and are predicted to be pathogenic by at least two dbNSFP tools (https://sites.google.com/site/jpopgen/dbNSFP) (**Table 1A**) (Liu et al., 2020).

Additionally, they flank the zinc finger domain of PRDM1 and may result in a loss of phosphorylation at these sites or affect protein folding and its ability to bind DNA (**Fig. 1B**) (Keller & Maniatis, 1992). Together, our data suggest that variants in *PRDM1* may result in SHFM phenotypes.

### Loss of Prdm1a causes pectoral fin defects

PRDM1 is required for vertebrate limb development (Ha & Riddle, 2003; Lee & Roy, 2006; Mercader et al., 2006; Robertson et al., 2007; Vincent et al., 2005; Wilm & Solnica-Krezel, 2005). The zebrafish pectoral fin is homologous to mammalian forelimbs, and its early structures consist of a cleithrum, scapulocoracoid/postcoracoid, endoskeletal disk, and fin fold, which are all derived from mesenchymal cells (**Fig. 2A**) (Grandel & Schulte-Merker, 1998). In zebrafish, *prdm1a* is first expressed in the pectoral fin at 18 hpf. It is highly expressed in the fin mesenchyme, pharyngeal arches, and neurons at 24 hpf (**Fig. 2B**). At 48 hpf, expression continues in the pharyngeal arches and neurons, and *prdm1a* is present in the apical ectodermal ridge (AER), the primary outgrowth signaling center of the limb (Wilm & Solnica-Krezel, 2005) (**Fig. 2C**). Previous studies have shown that knockdown of *prdm1a* using anti-sense morpholinos completely disrupts pectoral fin growth (Mercader et al., 2006). Hypomorph *prdm1a*^*tp39/tp39*^ mutants, which have a missense mutation (p.H564R) in the second zinc finger (**Fig. 2D**), present with mild phenotypes, namely a shortening of the scapulocoracoid and variable truncation of the fin, which is incompletely penetrant, occurring in only 30% of mutants (Baxendale et al., 2004; Lee & Roy, 2006; Roy et al., 2001). Predicted null *prdm1a*^*m805/m805*^ mutants, hereafter referred to as *prdm1a*^*-/-*^, have a mutation resulting in a premature stop codon in the SET domain (p.W154*) (**Fig. 2D**) (Artinger et al., 1999; Hernandez-Lagunas et al., 2005). To evaluate fin phenotypes in *prdm1a*^*-/-*^ mutants, embryos were stained with Alcian blue to assess cartilage development at 4 dpf (days post fertilization). There are no significant differences between wildtype and *prdm1a*^*+/-*^ heterozygotes (**Fig. S1**). Heterozygotes were included in all wildtype measurements. *prdm1a*^*-/-*^ mutants present with severe pectoral fin defects (**Fig. 2E-J**). There is a significant decrease in the average length of the cleithrum (∼20% decrease, p=0.0173), endoskeletal disk (8.7%, p=0.0816), and fin fold (10.6%, p=0.0374) (**Fig. 2G-J**). These data suggest that the zinc finger domain is important for the function of Prdm1a in pectoral fin development and is specifically required for outgrowth of the skeletal elements.

**Figure 2.**
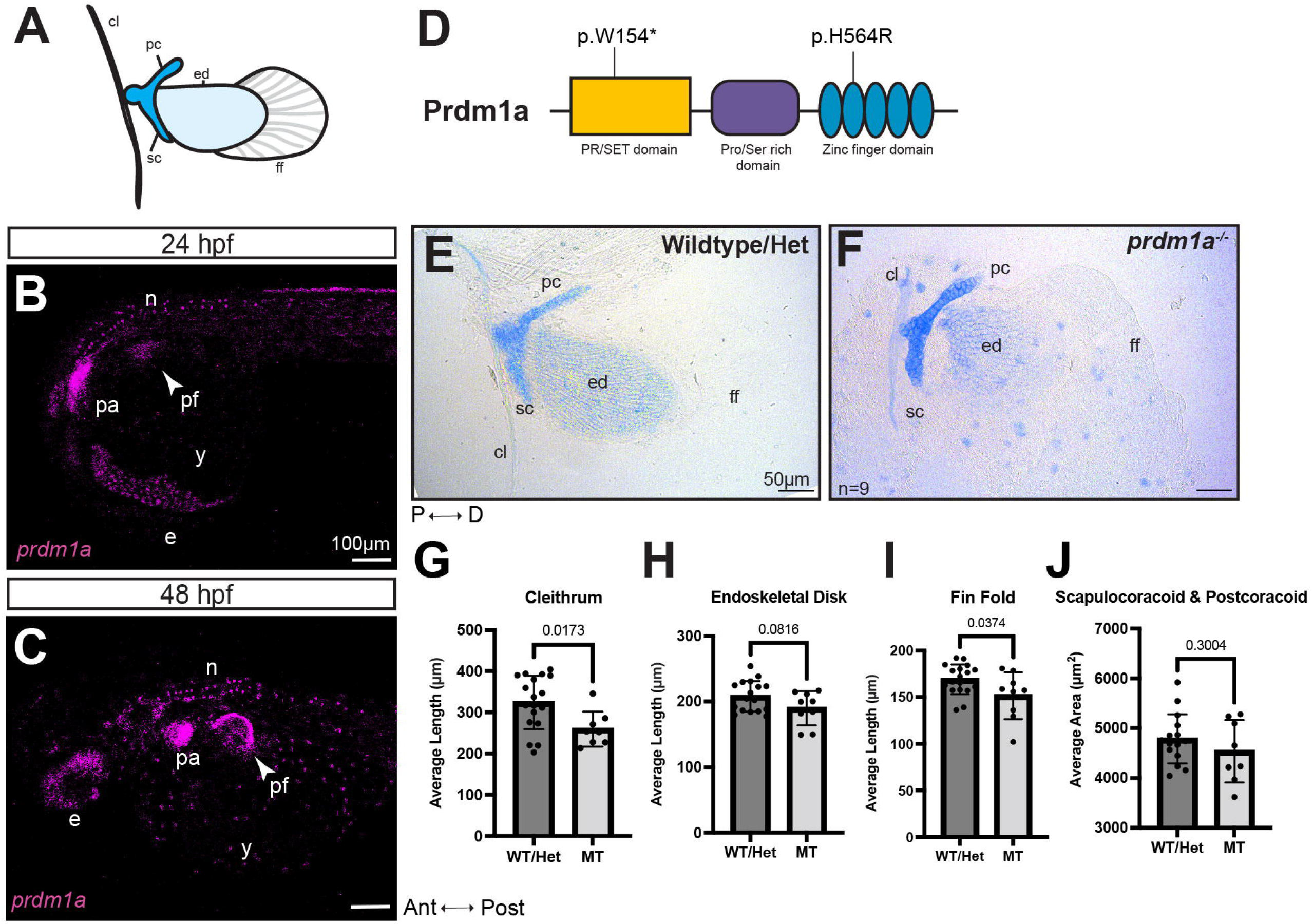
*prdm1a*^*-/-*^ mutants have hypoplastic pectoral fins. (**A**) Cartoon of pectoral fin bud at 4 dpf. (**B, C**) Lateral view of whole-mounted embryos after hybridization chain reaction (HCR) for *prdm1a* at (**B**) 24 (n=3) and (**C**) 48 hpf (n=28). Anterior is to the left. Images are 3D max projections of the whole embryo. Background was subtracted using the rolling ball feature in ImageJ (50 pixels). Arrows point to the pectoral fin bud. Scale bars are 100 µm. (**D**) Schematic of Prdm1a protein. The *prdm1a*^*m805/m805*^ allele causes a premature stop codon in the SET domain and is a presumed null (p.W154*). The hypomorphic *prdm1a*^*tp39/tp39*^ allele is a missense mutation in the second zinc finger (p.H564R). (**E, F**) Representative images of Alcian stained pectoral fins for (**E**) wildtype/heterozygous (n=18) and (**F**) *prdm1a*^*-/-*^ mutants (n=9) at 4 dpf. The average length of the (**G**) cleithrum, (**H**) endoskeletal disk, (**I**) fin fold, and (**J**) the average area of the scapulocoracoid/postcoracoid were measured. Averages were compared with an unpaired, two-tailed, independent student’s t-test. Error bars represent the mean ± SD. Scale bars are 50 µm. Abbreviations: ant, anterior; cl, cleithrum; d, distal; e, eye; ed, endoskeletal disk; ff, fin fold; het, heterozygous; hpf, hours post fertilization; MT, *prdm1a*^*-/-*^ mutant; n, neurons; p, proximal; pc, postcoracoid; pf, pectoral fin; post, posterior; sc, scapulocoracoid; WT, wildtype; y, yolk

### SHFM *hPRDM1* variants are not functional

To test whether the SHFM *hPRDM1* variants are functional, we designed an *in vivo* pectoral fin rescue experiment, where *hPRDM1* variants were overexpressed. We overexpressed either wildtype *hPRDM1* mRNA or each of the three SHFM variants in intercrossed *prdm1a*^*+/-*^ zebrafish embryos. In *prdm1a*^*-/-*^ mutants, injection of wildtype *hPRDM1* partially rescues the pectoral fin and more closely resembles those of uninjected wildtype (**Fig. 3A-C**), particularly the length of the cleithrum (12% increase), endoskeletal disk (5.5%), and fin fold (6%) (**Fig. 3G-J**). Inability of the wildtype allele to fully rescue the pectoral fin is likely due to the transience of the assay and rapid degradation of the mRNA. In contrast, overexpression of the three SHFM variants fails to rescue the elements of the pectoral fin (**Fig. 3D-F**). Indeed, injection of *hPRDM1* variant (p.T819A) in mutants further exacerbates hypoplasia of the endoskeletal disk (p=0.0138) (**Fig. 3F,H**). These data suggest that the SHFM variants are pathogenic and have reduced function compared to the wildtype allele. Interestingly, overexpression of the SHFM variants *hPRDM1* (p.C239Lfs*32) and (p.T819A) led to a significant decrease in endoskeletal disk length in wildtype embryos (p=0.0135 and p=0.0280, respectively), suggesting a dominant negative effect of the alleles on pectoral fin development (**Fig. S2**). Given the location of the alleles, we would predict that the zinc finger domain is important for the function of PRDM1 in limb development.

**Figure 3.**
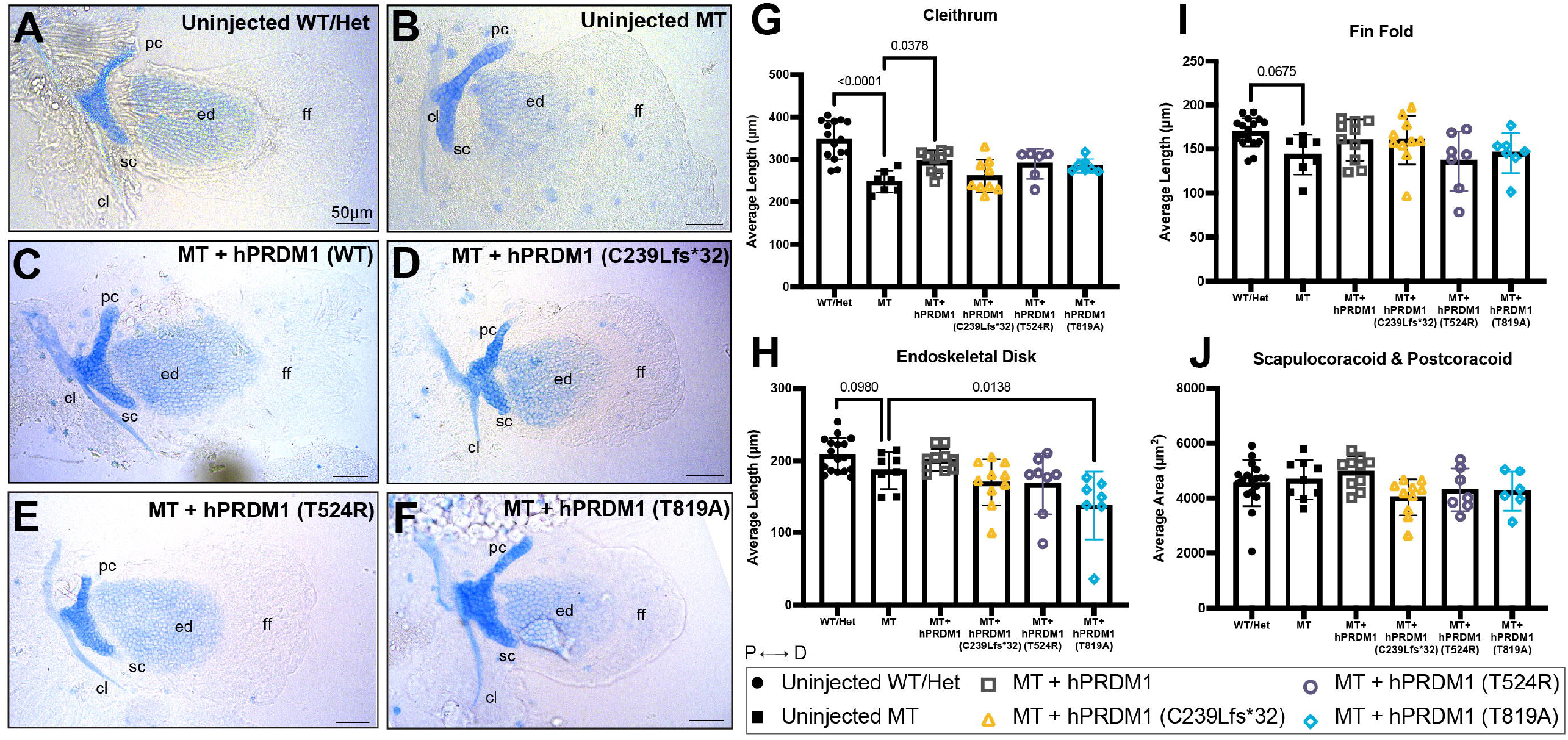
Transient overexpression of SHFM *hPRDM1* variants fails to rescue pectoral fin in *prdm1a*^*-/-*^ mutants. *prdm1a*^*+/-*^ heterozygous fish were intercrossed and injected with the *hPRDM1* wildtype and SHFM variant mRNA at the single-cell stage. Injected larvae were collected at 4 dpf for Alcian blue staining. **(A-F**) Representative images of Alcian stained pectoral fins at 4 dpf. (**A**) Uninjected wildtype/heterozygous (n=18). (**B**) Uninjected *prdm1a*^*-/-*^ mutant (n=9). *prdm1a*^*-/-*^ mutants were injected with (**C**) wildtype *hPRDM1* (n=10), (**D**) *hPRDM1*(p.C239Lfs*32) (n=10), (**E**) *hPRDM1*(p.T524R) (n=8), or *hPRDM1*(p.T819A) mRNA (n=7). Measurements for the length of the (**G**) cleithrum, (**H**) endoskeletal disk, and (**I**) fin fold and (**J**) the area of the scapulocoracoid and postcoracoid were averaged and compared using a one-way ANOVA, followed by a Tukey post-hoc test relative to uninjected *prdm1a*^*-/-*^ mutants. Error bars represent the mean ± SD. Scale bars are 50 µm. Abbreviations: cl, cleithrum; d, distal; ed, endoskeletal disk; ff, fin fold; het, heterozygous; hpf, hours post fertilization; MT, *prdm1a*^*-/-*^ mutant; p, proximal; pc, postcoracoid; sc, scapulocoracoid; WT, wildtype

### Prdm1a proline/serine-rich and DNA-binding zinc finger domains are required to regulate pectoral fin development

PRDM1 has an N-terminal SET domain, followed by a pro/ser-rich domain, and five zinc fingers. Although it lacks histone methyltransferase activity at its SET domain, PRDM1 is capable of modifying chromatin by recruiting epigenetic modifiers to either its pro/ser-rich or zinc finger domains (Ancelin et al., 2006; Gyory et al., 2004; Prajapati et al., 2019; Su et al., 2009; Yu et al., 2000). It can also recruit Groucho proteins as co-repressors to its pro/ser domain (Ren et al., 1999) or bind directly to DNA at its zinc finger domain to activate transcription (Powell et al., 2013; von Hofsten et al., 2008).

Moreover, the PRDM family of proteins regulate transcription in a highly cell/tissue specific manner (Fog et al., 2011).

To determine the functionally active domain of PRDM1 during limb development, we overexpressed modified versions of Prdm1a in null mutants using a stable, conditional Gal4/UAS system (**Fig. 4A-B**). Transgenic fish expressing Gal4 under a heat shock promoter, *Tg(hsp70l:gal4)*^*co1025Tg*^, were generated and crossed to *prdm1a*^*+/-*^ to create *Tg(hsp70I:gal4);prdm1a*^*+/-*^ fish. Using site-directed mutagenesis, we deleted each of the three functional domains of Prdm1a (**Fig. 4A; Table S2**). These deletions were modeled after previous *in vitro* studies (Gyory et al., 2004; Ren et al., 1999; Su et al., 2009; Yu et al., 2000). The modified genes were tagged with a self-cleaving 2a-EGFP reporter and placed under the control of a 4Xnr *UAS* enhancer. At the single cell-stage, we injected the *4XnrUAS-modified prdm1a-2a-EGFP* constructs into *Tg(hsp70l:gal4);prdm1a*^*+/-*^ intercrossed embryos along with Tol2 transposase mRNA. During normal development, *prdm1a* is first expressed in the pectoral fin at 18 hpf.

**Figure 4.**
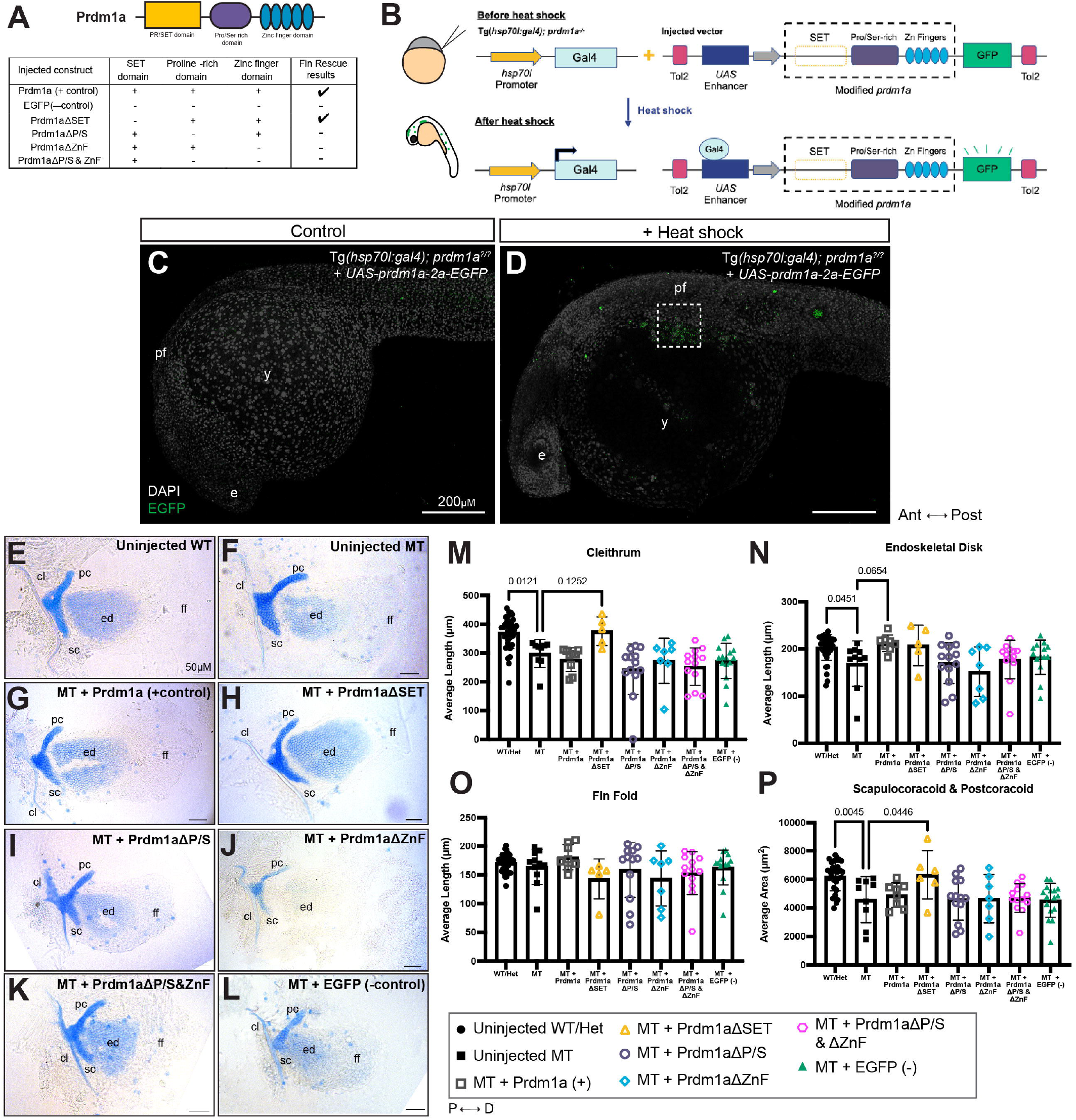
Overexpression of Prdm1a using a global heat shock Gal4/UAS system shows proline/serine-rich and zinc finger domains are required for pectoral fin function. (**A**) Schematic of *4XnrUAS-modified prdm1a-2a-EGFP* constructs that were injected into *Tg(hsp70l:gal4FF);prdm1a*^*+/-*^ intercrosses. Results for the ability to rescue the pectoral fin are shown. (**B**) Experimental design for heat shock Gal4/UAS rescue experiments. Following injection with the *UAS* construct, embryos at 6 hpf (shield stage) are heat shocked, leading to activation of Gal4, expression of the *4XnrUAS-modified prdm1a-2a-EGFP* construct, and cleavage of the 2a viral peptide from EGFP. Embryos were screened for mosaic EGFP expression at 24 hpf. (**C-D**) Representative images of 24 hpf embryos injected with the *4XnrUAS-modified prdm1a-2a-EGFP* construct at the single-cell stage. (**C**) No heat shock (control). (**D**) Mosaic EGFP expression in embryos injected and heat shocked. The dotted line marks the pectoral fin. Scale bars are 200 µm. (**E-K**) Representative images of Alcian stained pectoral fins at 4 dpf are shown. (**E**) Uninjected wildtype (n=36). (**F**) Uninjected *prdm1a*^*-/-*^ mutants (n=11). Mutants were injected with constructs containing (**G**) full-length Prdm1a (n=9), (**H**) Prdm1aΔSET (n=7), (**I**) Prdm1aΔP/S (n=13), (**J**) Prdm1aΔZnf (n=7), (**K**) Prdm1aΔP/S&Znf (n=13), and (**L**) an EGFP negative control (n=16). Scale bars are 50 µm. Measurements were taken for the length of the (**M**) cleithrum, (**N**) endoskeletal disk, and (**O**) fin fold and (**P**) the area of the scapulocoracoid and postcoracoid. Measurements for each individual were averaged and compared using a one-way ANOVA, followed by a Tukey’s post-hoc test relative to uninjected, heat shocked *prdm1a*^*-/-*^ mutants. Error bars represent the mean ± SD. Abbreviations: Δ, deleted; ant, anterior; cl, cleithrum; d, distal; dpf, days post fertilization; e, eye; ed, endoskeletal disk; ff, fin fold; hpf, hours post fertilization; MT, *prdm1a*^*-/-*^ mutant; p, proximal; pc, postcoracoid; pf, pectoral fin; post, posterior; sc, scapulocoracoid; WT, wildtype; y, yolk

Therefore, we heat shocked the embryos at 6 hpf (shield stage), giving the embryos time to transcribe, translate, and activate the Gal4 protein. Gal4 protein then binds to UAS and activates transcription of the modified *prdm1a* construct. At 24 hpf, we screened embryos for mosaic EGFP expression (**Fig. 4C-D**), and then stained them with Alcian blue at 4 dpf to assess the level of rescue to the pectoral fin compared to uninjected controls (**Fig. 4E-L; Fig. S3**). Of note, mosaic EGFP expression in injected and heat shocked embryos was highly variable and may have played a role in the construct’s ability to rescue the pectoral fin. *prdm1a*^*-/-*^ mutants injected with the positive control, full-length Prdm1a, exhibit a rescue, particularly in the endoskeletal disk (25.6% increase, p=0.0654) compared to uninjected controls (**Fig. 4F,G,N**). Interestingly, deletion of the SET domain (Prdm1aΔSET) can also partially rescue the area of the scapulocoracoid/postcoracoid (38.0% increase, p=0.0446) (**Fig. 4H,P**), cleithrum (25.8%, p= 0.1252) (**Fig. 4M**) and endoskeletal disk (22.9%) (**Fig. 4N**), suggesting that this domain is not important for pectoral fin development. However, when either the proline/serine (Prdm1aΔP/S) or zinc finger (Prdm1aΔZnF) domain is deleted, the construct fails to rescue the pectoral fin (**Fig. 4I-J, M-P**). Furthermore, when the two domains are deleted together (Prdm1aΔP/S&ZnF), we fail to see a rescue in any of the structures (**Fig. 4K, M-P**). The pro/ser and zinc finger domains are important for recruiting epigenetic modifiers and binding DNA, respectively. Injection of a negative EGFP control also fails to rescue any cartilage structures in the pectoral fin of *prdm1a*^*-/-*^ (**Fig. 4L-P**). Our data suggest that Prdm1a requires both its proline/serine and zinc finger domains to properly regulate fin development.

### Prdm1a controls Fgf signaling in the fin mesenchyme and maintenance of outgrowth and patterning genes

To determine the molecular mechanism by which Prdm1a regulates pectoral fin development, transgenic fish expressing EGFP under a mouse *Prx1* enhancer, *Tg*(*MmuPrx1:EGFP*)^*co1026Tg*^, were generated and crossed to *prdm1a*^*+/-*^ to create *Tg(MmuPrx1:EGFP;prdm1a*^*+/-*^ heterozygous fish. At 48 hpf, this transgene labels the pectoral fin, pharyngeal arches, and dorsal part of the head with EGFP (**Fig. 5A, A’**) (Hernandez-Vega & Minguillon, 2011; Yano & Tamura, 2013).

**Figure 5.**
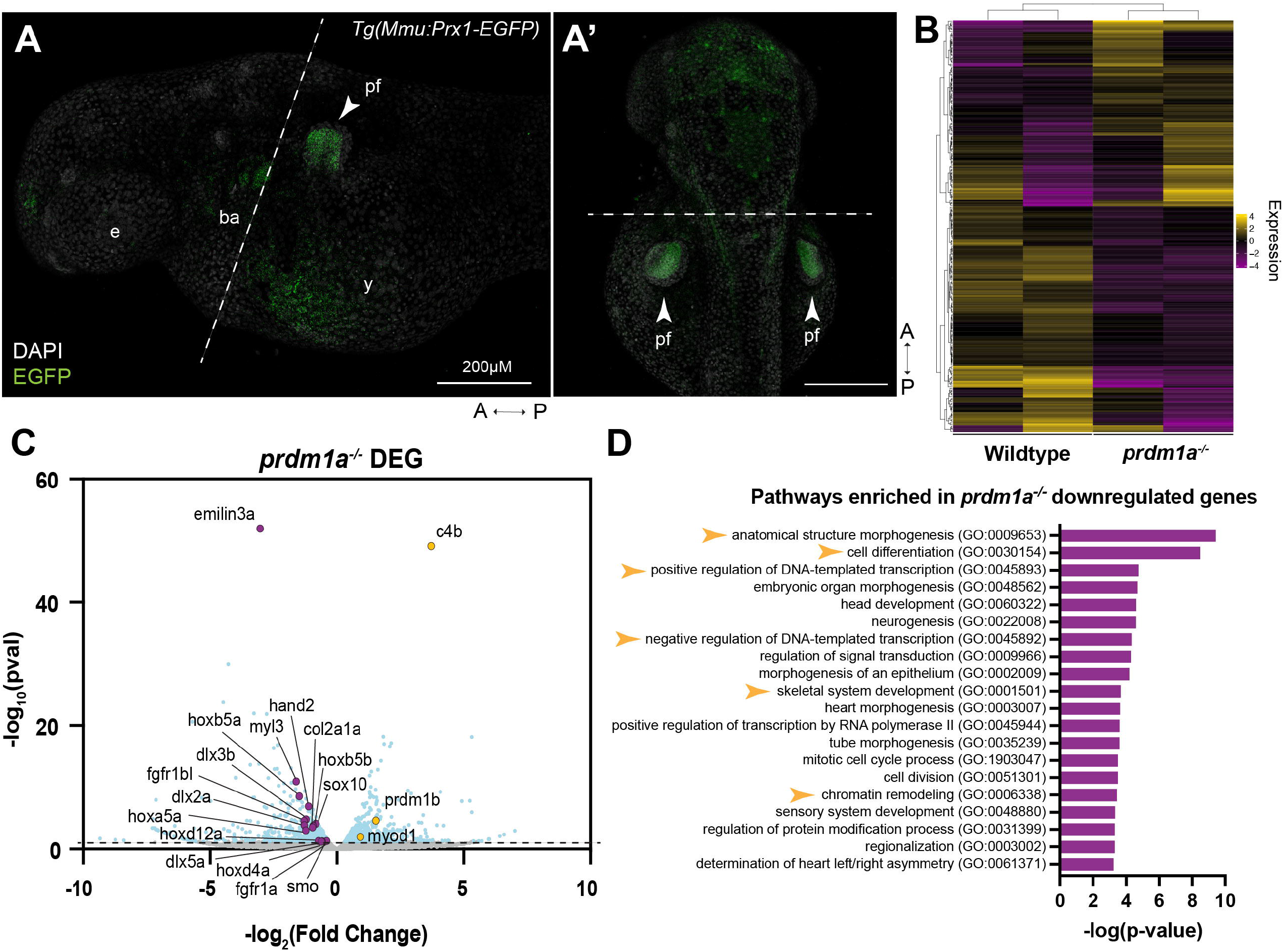
Loss of Prdm1a leads to downregulation of important limb development genes in the pectoral fin. RNA-seq was performed on isolated pectoral fin cells from wildtype and *prdm1a*^*-/-*^ embryos at 48 hpf. (**A**) Lateral and (**A’**) dorsal view of EGFP-positive pectoral fins from *Tg(MmuPrx1-EGFP)* zebrafish line at 48 hpf before FAC sorting. Dotted lines indicate where the embryos were dissected prior to the FAC sort. Scale bars are 200 µm. (**B**) Heat map of top 250 differentially expressed genes (padj) between wildtype and *prdm1a*^*-/-*^. (**C**) Volcano plot showing spread of differentially genes in pectoral fins of *prdm1a*^*-/-*^ compared to wildtype. Light blue dots are significant, differentially expressed genes (-log_10_ p-value≥1.15). Purple dots are downregulated, selected genes of interest, while yellow dots are upregulated. (**D**) Downregulated genes in *prdm1a*^*-/-*^ were subjected to GO (Panther) Pathway Enrichment Analysis. Yellow arrows highlight pathways of interest. Abbreviations: a, anterior; DEG, differentially expressed genes; e, eye; p, posterior; pf, pectoral fin; y, yolk

*Tg(Mmu:Prx1:EGFP);prdm1a*^*+/-*^ embryos were intercrossed, and at 48 hpf, wildtype and *prdm1a*^*-/-*^ embryos were dissected to remove the head and pharyngeal arches. EGFP*-* positive pectoral fin cells were isolated using fluorescence-activated cell (FAC) sorting before they were subjected to bulk RNA-sequencing (RNA-seq) on the Illumina NovaSEQ 6000 system. Our RNA-seq analysis revealed a total of 1,476 differentially expressed genes between wildtype and *prdm1a*^*-/-*^ mutants specifically in the pectoral fin (-log_10_≥1.2). Of these, 768 were upregulated, while 708 were downregulated (**Fig. 5B-C**). The most significant downregulated gene was *emilin3a*, a glycoprotein within the extracellular matrix belonging to the EMILIN/multimerin family, that to date has been shown to be expressed in the notochord, pharyngeal arches, and developing craniofacial skeleton of zebrafish (Corallo et al., 2013; Milanetto et al., 2007). The *Tg(Mmu:Prx1-EGFP)* transgenic line used for the RNA-seq also expresses EGFP in the pharyngeal arches and dorsal part of the head at 48 hpf (Hernandez-Vega & Minguillon, 2011; Yano & Tamura, 2013). Though we dissected and removed these regions prior to FAC sorting, there may have been some residual arch and dorsal head tissue in our samples. Key limb genes known to be involved in pectoral fin development, including members of the *hoxa* and *hoxd* gene families, *dlx2a, dlx5a, hand2, col2a1a, smo*, and *fgfr1a/fgfr1bl*, are significantly downregulated in *prdm1a*^*-/-*^ (**Fig. 5C**), which are required for induction, patterning, outgrowth, and collagen production (Ahn & Ho, 2008; Akimenko et al., 1994; Chen et al., 2001; Dale & Topczewski, 2011; Heude et al., 2014; Leerberg et al., 2019; Yan et al., 1995; Yelon et al., 2000). The most significant upregulated gene was complement factor 4b (c4b), which is part of the classical activation pathway in the immune system (Janeway et al., 2001). The paralog *prdm1b* was also upregulated in *prdm1a*^*-/-*^, suggesting an attempt at genetic compensation.

Gene ontology (GO) pathway enrichment analysis on genes downregulated in *prdm1a*^*-/-*^ revealed anatomical structure morphogenesis, chromatin remodeling, skeletal system development, cell differentiation, and transcriptional regulation were among the pathways most enriched in the downregulated genes (**Fig. 5D**). These pathways were expected given what we already know about PRDM1 as a transcription factor and master regulator of differentiation (reviewed in Bikoff et al. (2009)).

To validate the RNA-seq data and determine the effect of *prdm1a* loss on gene expression, we performed real-time quantitative PCR (RT-qPCR) on the anterior half of embryos at 24 and 48 hpf and hybridization chain reaction (HCR) at 48 hpf for select genes in wildtype and *prdm1a*^*-/-*^ whole embryos. (**Fig. 6; Fig. S4**). We first probed for *prdm1a* and *tbx5a*, an early marker of limb initiation. At 48 hpf, *prdm1a* is highly expressed in the fin mesenchyme and AER of wildtype embryos. In null mutants, *prdm1a* transcripts are detected throughout the fin bud and at even higher levels than wildtype, suggesting that the mRNA is not susceptible to nonsense mediated decay at this stage (**Fig. 6**). The cells may be overproducing *prdm1a* transcripts to compensate for the loss of functional protein. *tbx5a* is highly expressed in the fin mesenchyme of both wildtype and *prdm1a*^*-/-*^ at 48 hpf with no significant difference (**Fig. 6A-B**). The intensity of signal in the fin mesenchyme was quantified by measuring the total cell fluorescence and correcting for area and background (CTCF). Next, Fgf receptors, *fgfr1a* and *fgfr1bl*, were significantly downregulated in the RNA-seq dataset (**Fig. 5C**), so we looked at the expression of their ligand, *fgf10a*, which has been shown to be decreased in hypomorphs and morphants (Lee & Roy, 2006; Mercader et al., 2006). In wildtype embryos, *fgf10a* is highly expressed in the fin mesenchyme, but this is significantly reduced in *prdm1a*^*-/-*^ (**Fig. 6C-G; Fig. S4A, G**). This was quantified by measuring the signal intensity along a line drawn from the most proximal to distal point of the fin bud and normalizing the intensity and distance between 0 and 1 (Fulton et al., 2020). FGF10 is a marker of limb induction and is known for signaling downstream to FGF8 in the AER to regulate outgrowth along the proximal distal axis (Ng et al., 2002; Ohuchi et al., 1997). In *prdm1a*^*-/-*^ embryos, *fgf8a* expression is significantly decreased, which is consistent with the observed truncated fin phenotype (**Fig. 6H-L Fig. S4L**).

**Figure 6.**
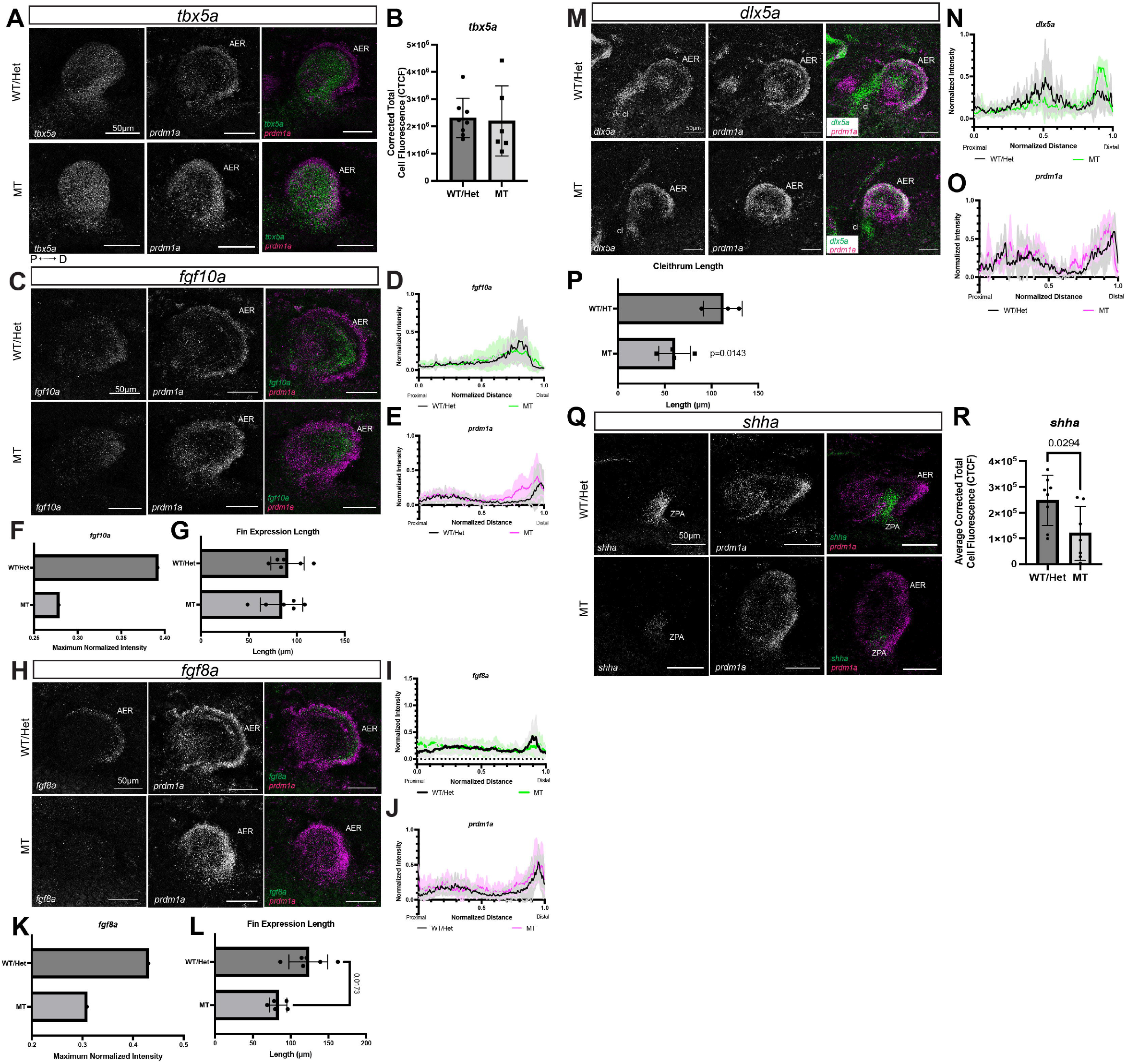
Prdm1a acts downstream of limb initiation and regulates FGF signaling in the fin mesenchyme required for outgrowth and anterior/posterior patterning. **A, C, H, M, Q**) Lateral views of pectoral fins from whole-mount wildtype and *prdm1a*^*-/-*^ mutant embryos after hybridization reaction (HCR) was performed at 48 hpf. (**A**) *tbx5a* (limb initiation) and *prdm1a* expression (n=8 WT, n=6 *prdm1a*^*-/-*^). (**B**) Quantification of *tbx5a* expression using corrected total cell fluorescence (CTCF). (**C**) *fgf10a* (limb induction) and *prdm1a* expression (n=6 for each genotype). (**D-E**) Quantification of (**D**) *fgf10a* and (**E**) *prdm1a* expression along a line drawn from the most proximal to the most distal point of the fin bud using the line scan tool on ImageJ. Intensity and distance were normalized between 0 and 1. (**F**) Maximum normalized intensity of *fgf10a*. (**G**) Length of the fin as measured by *fgf10a* gene expression. (**H**) *fgf8a* (AER outgrowth marker) and *prdm1a* expression (n=6 for each genotype). (**I-J**) Quantification of (**I**) *fgf8a* and (**J**) *prdm1a* expression along a line drawn from the most proximal to the most distal point of the fin bud. (**K**) Maximum normalized intensity of *fgf8a*. (**L**) Length of the fin as measured by *fgf8a* gene expression. (**M**) *dlx5a* (outgrowth marker) and *prdm1a* expression (n=3 WT, n=4 *prdm1a*^*-/-*^). (**N-O**) Quantification of (**N**) *dlx5a* and (**O**) *prdm1a* expression along a line drawn from the most proximal to the most distal point of the fin bud. (**P**) Length of the cleithrum. (**Q**) *shha* (anterior/posterior patterning) and *prdm1a* expression (n=8 for each genotype). (**R**) Expression of *shha* was quantified using CTCF. Scale bars are 50 µm. Solid lines in line intensity graphs represent the mean ± SD. Statistical comparisons were made using unpaired, two-tailed, independent student’s t-test. All images are 3D max projections of lateral views of the pectoral fin. Background was subtracted using the rolling ball feature in ImageJ (50 pixels). Abbreviations: AER, apical ectodermal ridge; CTCF, corrected total cell fluorescence; d, distal; hpf, hours post fertilization; p, proximal; ZPA, zone of polarizing activity

RNA-seq also shows that *dlx5a*, another marker of limb outgrowth, is downregulated (**Fig. 5C**). By HCR, *dlx5a* is highly expressed in the fin mesenchyme and cleithrum and is co-expressed with *prdm1a* in AER cells (**Fig. 6M**). Intriguingly, *dlx5a* expression is decreased in the mesenchyme and cleithrum of *prdm1a*^*-/-*^ but increased in the AER of *prdm1a*^*-/-*^ (**Fig. 6N-P; S4D, J**). Finally, we probed for sonic hedgehog (*shha*), the morphogen required for anterior/posterior digit patterning. In the most posterior part of the fin bud, the zone of polarizing activity (ZPA), there is a significant decrease of *shha* in *prdm1a*^*-/-*^ (**Fig. 6Q-R)**. This was expected in that we also see a downregulation of the receptor and SHH target *smo* in our RNA-seq dataset (**Fig. 5C**). Our gene expression results in null *prdm1a*^*-/-*^ mutants are consistent with published studies in mice as well as morphant and hypomorph *prdm1a*^*tp39/tp39*^ zebrafish studies (Lee & Roy, 2006; Mercader et al., 2006; Robertson et al., 2007). Together, these data suggest that Prdm1a is required for regulating FGF signaling in the fin mesenchyme as well as outgrowth and anterior/posterior patterning in the AER and ZPA.

### Prdm1a directly binds to and regulates limb outgrowth genes

Given that Prdm1a requires its zinc finger domain during pectoral fin development, we next asked whether Prdm1a directly binds to limb genes that were identified in the RNA-seq to regulate their expression. We isolated EGFP-positive pectoral fin cells from *Tg(Mmu:Prx1-EGFP)* wildtype fish at 24 hpf (**Fig. 7A**) and performed Cleavage Under Targets and Release Using Nuclease (CUT&RUN).

**Figure 7.**
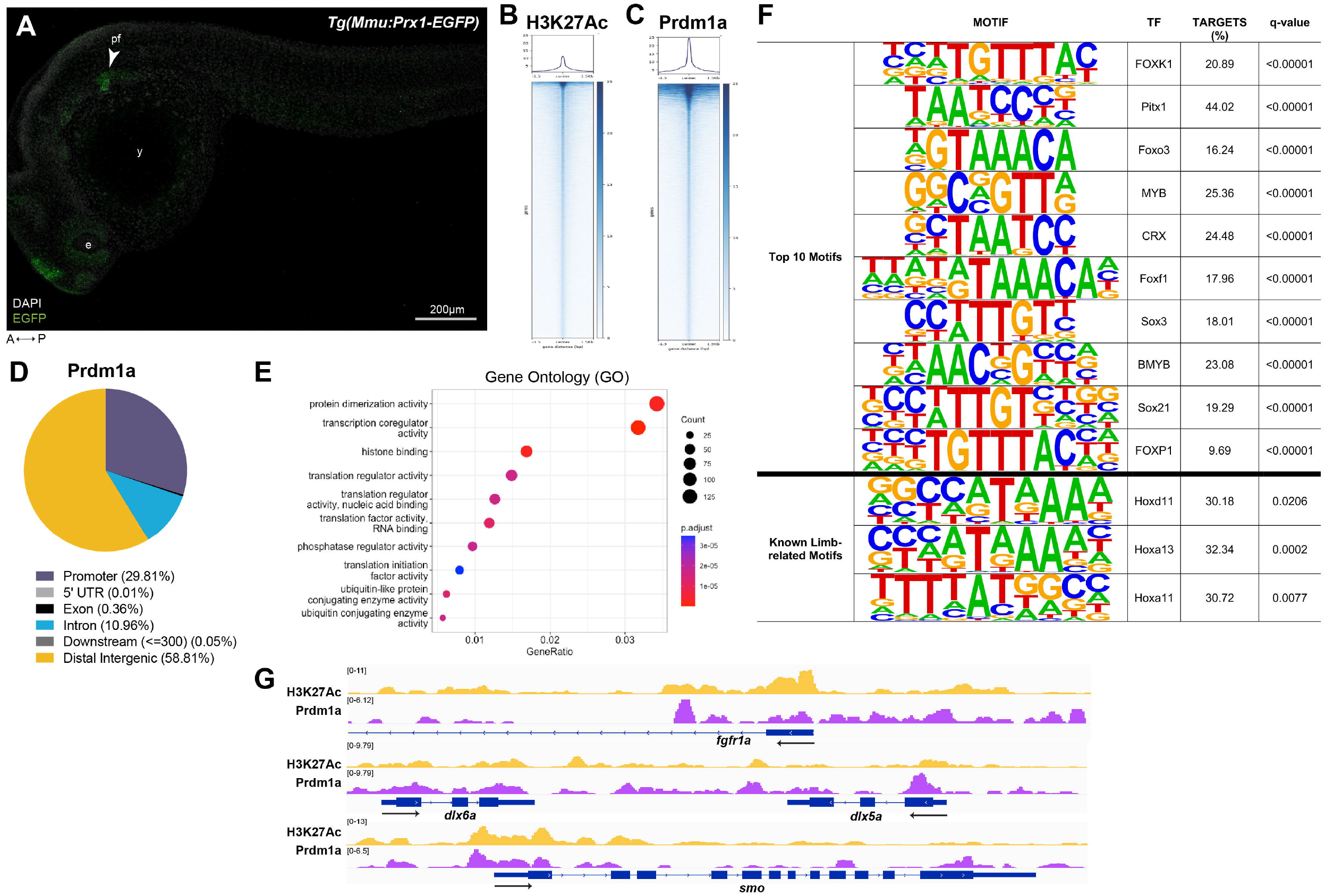
Prdm1a directly binds to and regulates limb genes. (**A**) Lateral view of EGFP-positive pectoral fins from Tg(*MmuPrx1-EGFP*) zebrafish line at 24 hpf before CUT&RUN was performed. Scale bar is 200 µm. (**B-C**) Coverage heatmaps of (**B**) H3K27Ac and (**C**) Prdm1a binding across the genome 1.5 kb upstream and downstream of the peak center. (**D**) Annotation of enriched binding sites by Prdm1a. (**E**) Enriched Prdm1a peaks were subjected to gene ontology (GO) terms analysis using ChIPseeker’s seq2gene function. (**F**) Prdm1a peaks were subjected to motif analysis using HOMER. The top 10 motifs as well as known limb-related motifs are shown. (**G**) Tracks showing H3K27Ac enrichment (open chromatin) and Prdm1a binding sites for *fgfr1a, dlx5a/dlx6a*, and *smo*. There is variability between replicates, but the overall trends are comparable (see **Fig. S5**). Abbreviations: e, eye; pf, pectoral fin; y, yolk

CUT&RUN is a high-resolution, *in situ* method for chromatin profiling and mapping of protein/DNA interactions, which uses fewer cells than traditional Chromatin Immunoprecipitation (ChIP) (Meers et al., 2019; Skene & Henikoff, 2017; Ye et al., 2021). We used antibodies against IgG, H3K27Ac, and Prdm1a (von Hofsten et al., 2008) and sequenced the samples on the Illumina NovaSEQ 6000 system. We observed 15,361 Prdm1a-occupied peaks (**Fig. 7B-C; Fig. S5A-B**). Of these, 29.81% were associated with promoter regions, 10.96% in introns, and 58.81% in distal intergenic regions (**Fig. 7D; Fig. S5C**). We then subjected the Prdm1a peaks to Gene ontology (GO) pathway analysis and found enrichment for pathways involved in transcriptional regulation, such as protein dimerization activity, transcription coregulator activity, and histone binding (**Fig. 7E**). We also performed motif enrichment analysis and identified a significant enrichment of Hox transcription factor binding sites. Several Hox transcription factors are known to be required for pectoral fin development, such as Hoxd11, Hoxa13, and Hoxa11 (**Fig. 7F; Fig. S5E; Table S3**) (Sordino et al., 1996; Sordino et al., 1995). To our knowledge, Prdm1a has not yet been shown to interact with Hox transcription factors. We also mapped Prdm1a binding sites to the nearest genes and found that Prdm1a directly binds to putative enhancer and promoter regions of critical limb development genes, including *fgfr1a, dlx5a, dlx6a*, and *smo* (**Fig. 7G; Fig. S5F**). This is consistent with what is observed in our RNA-seq dataset in that the expression of these genes is significantly downregulated in *prdm1a*^*-/-*^ compared to wildtype embryos. The data suggest that Prdm1a directly binds to these genes and functions as an activator to regulate limb induction, outgrowth, and anterior/posterior patterning.

## DISCUSSION

Approximately 50% of SHFM cases have an unknown genetic cause.

Chromosomal deletions and translocations at 6q21 have long been associated with SHFM, though a candidate gene has not yet been isolated (Braverman et al., 1993; Correa-Cerro et al., 1996; Duran-Gonzalez et al., 2007; Gurrieri et al., 1995; Hopkin et al., 1997; Pandya et al., 1995; Tsukahara et al., 1997; Viljoen & Smart, 1993). In this study, we identified three novel variants in the gene *PRDM1* in families with SHFM that segregated with the phenotype or arose *de novo*. PRDM1 has been previously implicated in vertebrate limb development (Ha & Riddle, 2003; Lee & Roy, 2006; Mercader et al., 2006; Robertson et al., 2007; Vincent et al., 2005; Wilm & Solnica-Krezel, 2005), and here we show that *PRDM1* variants can cause SHFM and limb defects in humans. Each of the three variants is rare, negatively affects the DNA-binding zinc finger domain of the protein and are damaging as heterozygous alleles. We have shown through transient overexpression assays in zebrafish that the variants are not functional compared to wildtype in that they fail to rescue cartilage elements of the pectoral fins of *prdm1a*^*-/-*^ zebrafish embryos. Our data suggest that they are pathogenic and led to limb defects in SHFM patients.

Using stable, conditional overexpression experiments, we also define the functional domain of Prdm1a specifically during limb development. PRDM1 consists of a SET domain at its N-terminus, followed by a pro/ser-rich domain, and five zinc fingers. PRDM1 can recruit epigenetic modifiers to its domains as well as bind DNA directly to regulate transcription. We show that Prdm1a requires both its pro/ser-rich and zinc finger domains for pectoral fin morphogenesis. Deleting either domain fails to rescue the pectoral fin, while deletion of the SET domain rescues the cleithrum, endoskeletal disk, scapulocoracoid and postcoracoid. The SET domain of PRDM1 does not have intrinsic methyltransferase activity *in vivo* and has not been shown to bind with cofactors (Cheng et al., 2005; Hohenauer & Moore, 2012; Martin & Zhang, 2005). Removing the pro/ser and zinc finger domains together also fails to rescue, but interestingly, it does not produce a more severe phenotype. This implies that both domains are required during pectoral fin development. Prdm1a likely directly binds to DNA with its zinc finger domain and then recruits cofactors to its pro/ser domain. If Prdm1a cannot bind, then neither can its cofactors. Likewise, binding to the DNA alone cannot repress or induce expression of that gene.

Within the limb GRN, Prdm1a has been proposed to act downstream of retinoic acid signaling and limb initiation and upstream of Fgf signaling to induce limb formation (Lee & Roy, 2006; Mercader et al., 2006). We performed RNA-seq on isolated pectoral fin cells and found that there was an almost equal distribution of up and downregulated genes in *prdm1a*^*-/-*^ compared to wildtype, though all key limb genes were downregulated (**Fig. 6B-C**). Given that Prdm1a is traditionally considered a gene repressor, it is surprising that the genes known to be involved in limb development were all downregulated in *prdm1a*^*-/-*^, including Fgf receptors, *col2a1a, dlx2a, dlx5a, smo*, and *hoxa/hoxd* genes (Ahn & Ho, 2008; Akimenko et al., 1994; Chen et al., 2001; Dale & Topczewski, 2011; Heude et al., 2014; Leerberg et al., 2019; Yan et al., 1995; Yelon et al., 2000). Using HCR, we demonstrate that *prdm1a*^*-/-*^ pectoral fins exhibit a significant decrease in important limb genes, namely *fgf10a, fgf8a, dlx5a*, and *shha*. We propose that during limb induction, Prdm1a promotes mesenchymal cell outgrowth and patterning by activating FGF receptor, *fgfr1a*. Binding of Fgf10a to this receptor then leads to downstream activation of *fgf8a* in the AER. Fgf8a initiates a positive feedback loop and maintains expression of *fgf10a* for sustained limb growth (Ng et al., 2002; Ohuchi et al., 1997). Fgf8a is also known to participate in initiation of SHH expression in the mesoderm and ZPA, which is responsible for anterior/posterior patterning (Crossley et al., 1996). Decreased expression of *fgf8a* subsequently leads to lower levels of *shha* in *prdm1a*^*-/-*^ (**Fig. 8**). Our data suggests that Prdm1a is necessary for initiating limb induction in the fin mesenchyme and maintenance of outgrowth and patterning genes.

**Figure 8.**
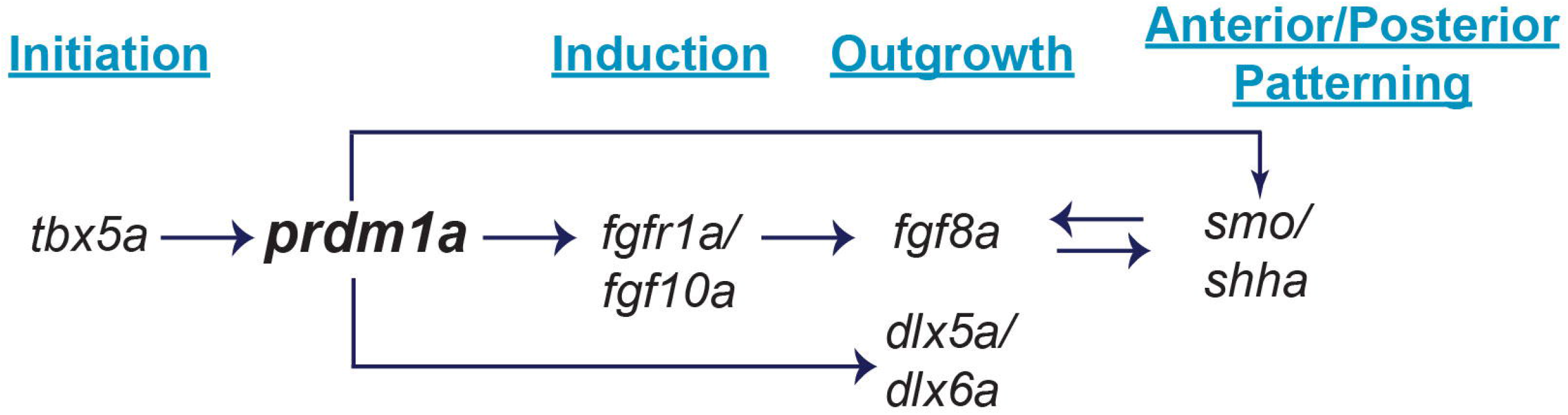
Working model of the Prdm1a gene regulatory network during pectoral fin development. Prdm1a acts downstream of limb initiation and *tbx5a* (**Fig. 6A**,**B**), but upstream of limb induction and Fgf signaling. Prdm1a directly binds to and activates *fgfr1a* (**Fig. 7G**), allowing Fgf10a to bind (**Fig. 6C-G**). Fgf10a then activates Fgf8a in the apical ectodermal ridge (**Fig. 6H-L**), signaling pectoral fin outgrowth. Fgf8a turns on Shh signaling in the zone of polarizing activity for anterior/posterior patterning (**Fig. 6Q-R**). Prdm1a also directly binds to *smo* (**Fig. 7G**), a receptor Shh signaling, as well as *dlx5a/*dlx6a, additional limb outgrowth markers (**Fig. 6M-P**).

Prdm1a has traditionally been known as a repressor, and it was initially thought that Prdm1a regulates Fgf signaling and the ensuing cascade by blocking the expression of an inhibitor of *fgf10a* transcription (Mercader et al., 2006). However, in this study, we show that Prdm1a directly binds to and regulates its receptor, *fgfr1a*, suggesting that it is *activating* Fgf signaling. We also show that Prdm1a directly binds to *smo* and activates anterior/posterior patterning. Its role as an activator is uncommon, though not novel. We have previously shown that Prdm1a directly activates genes, such as *tfap2* and *foxd3*, during zebrafish neural specification (Powell et al., 2013). More recently, it is has been shown to interact with Kdm4a, a histone demethylase, to activate chick neural, neural crest, and sensory specification genes (Prajapati et al., 2019).

Given that its traditional cofactors are repressors, i.e., HDAC1/2, Groucho proteins, LSD1, Prmt5 (Ancelin et al., 2006; Ren et al., 1999; Su et al., 2009; Yu et al., 2000), it is unlikely that Prdm1a is binding with these factors during the activation of these particular limb genes. However, “chromatin modification” was one of the most downregulated pathways in *prdm1a*^*-/-*^ from our GO Pathway Enrichment analysis of the RNA-seq dataset (**Fig. 5D**), and “transcription coregulator activity” was enriched in the CUT&RUN dataset (**Fig. 7E)**. Motif analysis of Prdm1a bound peaks predicts enrichment of Hox transcription factor motifs, including Hoxd11, Hoxa13, and Hoxa11, that are known to be required for pectoral fin development (**Fig. 7F**) (Sordino et al., 1996; Sordino et al., 1995). We also hypothesize that Prdm1a may be acting with Kdm4a to directly activate *fgfr1a* and *smo*, though additional experiments are needed to determine this or uncover other cofactors.

One of the more interesting genes to emerge from our transcriptomic and CUT&RUN analyses was *dlx5a*, a homeodomain transcription factor known to cause SHFM Type I (Crackower et al., 1996; Scherer et al., 1994; Wang et al., 2014). In our HCR assays we show that at 48 hpf, *prdm1a*^*-/-*^ exhibit decreased expression of *dlx5a* in the mesenchyme, but increased expression in the AER where *prdm1a* is also co-expressed (**Fig. 6M, N**). Given that *DLX5* mutations lead to SHFM Type I (MIM #225300) (Crackower et al., 1996; Scherer et al., 1994; Wang et al., 2014) and that *dlx5a* is directly regulated by Prdm1a, we asked whether there is a genetic interaction between the two genes. We crossed a hypomorphic allele *dlx5a*^*j1073Et*^ (referred to as *Tg(dlx5a:EGFP))* (Talbot et al., 2010) to *prdm1a*^*+/-*^ fish, intercrossed the double heterozygous animals, and performed cartilage staining on the resulting larvae at 4 dpf. *dlx5a* morphants have pectoral fin defects that vary in severity (Heude et al., 2014).

Interestingly, *Tg(dlx5a:EGFP)* heterozygotes and homozygotes do not have overt pectoral fin defects at this stage (**Fig. S6C, E**). When combined with *prdm1a*^*-/-*^, we found that the length of the endoskeletal disk and the area of the scapulocoracoid and postcoracoid were slightly increased and trending towards a rescue; however, the results were not significant (**Fig. S6D, F, H, J**). The partial loss of Dlx5a could have helped balance the high expression in the AER of *prdm1a*^*-/-*^ and rescue pectoral fin outgrowth, but, because it is a hypomorph, it may have been too weak to produce a more drastic rescue. In addition, expression of *dlx5a* in the cleithrum was significantly decreased in *prdm1a*^*-/-*^ (**Fig. 6M, O**). The cleithrum is part of the shoulder girdle in bony fishes. It is located at the border between the neural crest-derived pharyngeal arches, which give rise to the craniofacial skeleton, and the mesodermal pectoral fin. Because of its position, some have hypothesized that, like the clavicle in mammals, the cleithrum may be composed of both neural crest (NC) and mesodermal cells (Matsuoka et al., 2005). This could have important implications in human disease in that craniofacial and limb defects often co-occur, including in SHFM (reviewed in Gurrieri and Everman (2013); Truong and Artinger (2021)). Although there is currently no evidence that NC cells contribute to the cleithrum (Kague et al., 2012), NC cells have been labeled in gill pillar cells of zebrafish (Mongera et al., 2013) as well as the posterior gill arches of the little skate (*Leucoraja erinacea*) (Sleight & Gillis, 2020). The gill arches are hypothesized to give rise to paired fins in jawed vertebrates, implying a serial homology between the two structures (Gegenbaur, 1878; Sleight & Gillis, 2020). Given the importance of Prdm1a in the pharyngeal arches and now the pectoral fin, it is interesting to speculate whether the two structures are connected (Artinger et al., 1999; Birkholz et al., 2009; Ha & Riddle, 2003; Lee & Roy, 2006; Mercader et al., 2006; Robertson et al., 2007; Roy & Ng, 2004; Vincent et al., 2005; Wilm & Solnica-Krezel, 2005).

In summary, we have identified novel variants in *PRDM1* that result in SHFM phenotypes and limb defects in humans. Variants affecting the protein’s ability to recruit cofactors and bind to DNA are detrimental for proper limb formation. We show that Prdm1a directly binds to *fgfr1a, dlx5a, dlx6a*, and *smo* during limb development and its ability to do so is critical for proper outgrowth and patterning (**Fig. 8**). This work is important for our understanding of the limb GRN and underscores the pathogenicity of human variants in *PRDM1*.

## MATERIALS & METHODS

### Zebrafish husbandry

Zebrafish were maintained as previously described (Westerfield, 2000). The wildtype (WT) strain used was AB (ZIRC) and the mutant lines used were *prdm1a*^*m805*^ *(nrd;* referred to as *prdm1a*^*-/-*^) (Artinger et al., 1999; Hernandez-Lagunas et al., 2005) and *dlx5a*^*j1073Et*^ (referred to as *Tg(dlx5a:EGFP)*) (Talbot et al., 2010). Embryos were staged following published standards as described (Kimmel et al., 1995). All experiments were reviewed and approved by the Institutional Animal Care and Use Committee (IACUC) at the University of Colorado Denver Anschutz Medical Campus and conform to the NIH regulatory standards of care and treatment.

### SHFM Patient Ascertainment

SHFM individuals were seen in the clinic due to a history of non-syndromic limb and digit malformations. X-ray and pedigree analyses indicated a diagnosis of non-syndromic SHFM with dominant inheritance but variable penetrance for each. In addition to testing for common variants associated with SHFM, standard chromosomal karyotyping and microarray were performed but did not reveal any abnormalities. In SHFM family #1 (*PRDM1c*.*712_713insT*) DNA derived from whole blood was used to perform whole exome sequencing, which identified variants in the *PRDM1* gene.

Targeted sequencing for *PRDM1* was then performed on an additional 75 unrelated SHFM patients seen at the clinic, which identified two additional variants.

### DNA extraction, exome sequencing, and analysis

Exome sequencing was performed at the University of Washington Center for Mendelian Genomics (UW-CMG). Briefly, library construction and exome capture were done using an automated 96-well plate format (Perkin-Elmer Janus II). 500ng of genomic DNA was subjected to a series of shotgun library construction steps, including fragmentation through acoustic sonication (Covaris), end-polishing and A-tailing, ligation of sequencing adaptors, and PCR amplification with dual 8 bp barcodes for multiplexing. Libraries underwent exome capture using the Roche/Nimblegen SeqCap EZ v2.0 (∼36.5 MB target). Prior to sequencing, the library concentration was determined by fluorometric assay and molecular weight distributions verified on the Agilent Bioanalyzer (consistently 150 ± 15bp). Barcoded exome libraries were pooled using liquid handling robotics prior to clustering (Illumina cBot) and loading. Massively parallel sequencing-by-synthesis with fluorescently labeled, reversibly terminating nucleotides is carried out on the HiSeq sequencer. Variant detection and genotyping were performed using the HaplotypeCaller (HC) tool from GATK (3.7). Variant data for each sample were formatted (variant call format [VCF]) as “raw” calls that contain individual genotype data for one or multiple samples and flagged using the filtration walker (GATK) to mark sites that are of lower quality/false positives [e.g., low quality scores (Q50), allelic imbalance (ABHet 0.75), long homopolymer runs (HRun> 3), and/or low quality by depth (QD < 5)].

Coding variants were filtered by selecting variants that have <0.005 minor allele frequency (MAF) in any population in the Genome Aggregation Database (gnomAD). Further selection was performed by selecting rare single nucleotide variants (SNVs) that were considered damaging by at least two bioinformatics tools in dbNSFP (Liu et al., 2016). For indels, bioinformatics analysis was performed using MutationTaster (Schwarz et al., 2010).

### SHFM Patient Sanger sequencing

PRDM1 variants were confirmed by Sanger sequencing using BigDye Terminator v3.1 Cycle Sequencing kit on an ABI3100 automatic DNA Analyzer (Applied BioSystems) following manufacturer’s instructions. The alignment and analysis of the sequences were done using the DNASTAR program (Lasergene).

### Alcian blue cartilage staining

Zebrafish were stained for cartilage as previously described (Walker & Kimmel, 2007). In short, 4 dpf larvae were fixed in 2% paraformaldehyde (PFA) at room temperature for one hour. Larvae were then washed in 100mM Tris (pH 7.5)/10mM MgCl_2_ before rocking overnight at room temperature in Alcian blue stain (pH 7.5) (0.04% Alcian Blue, 80% ethanol, 100mM Tris [pH 7.5], 10mM MgCl_2_). Larvae were destained and rehydrated in a series of ethanol washes (80%, 50%, 25%) containing 100mM Tris (pH 7.5) and 10mM MgCl_2_, and then bleached for 10 minutes in 3% H_2_O_2_/0.5% KOH. Finally, larvae were rinsed twice in 25% glycerol/0.1% KOH to remove the bleach and stored at 4°C in 50% glycerol/0.1% KOH. The pectoral fins of stained larvae were dissected, flat mounted in 50% glycerol/0.1 KOH, and imaged on an Olympus BX51 WI microscope. Measurements of the pectoral fin were performed blindly in ImageJ, averaged for each individual, and then compared using a one-way ANOVA followed by a Tukey’s post-hoc test relative to uninjected *prdm1a*^*-/-*^ mutants.

### mRNA overexpression in zebrafish

*hPRDM1* variant cDNA was synthesized into a pCS2+ backbone using Gateway cloning. cDNA was linearized and transcribed using the mMessage mMachine T7 Transcription Kit (ThermoFisher). *prdm1a*^*+/-*^ fish were intercrossed and the different *hPRDM1* mRNA variants (diluted 1:10 in water and phenol red) were injected into resulting embryos at the single-cell stage. At 4 dpf, larvae were collected for Alcian blue staining.

### Hybridization chain reaction (HCR) V3.0 and quantification

Probes for *prdm1a, fgf10a, fgf8a, dlx5a, tbx5a*, and *shh* were purchased from Molecular Instruments (www.molecularinstruments.com). Whole mount HCR was performed according to the manufacturer’s instructions with minor modifications (Choi et al., 2016; Choi et al., 2018). Embryos were fixed overnight at 4°C in 4% PFA, washed in PBS, and dehydrated and permeabilized in two 10-minute washes in 100% MeOH at room temperature. Embryos were stored for at least 24 hours at -20°C in fresh MeOH. A graded series of MeOH/PBST solutions was used to rehydrate the embryos (75%, 50%, 25%, 0%). Embryos were then treated with proteinase K (10 μg/mL) for 5 minutes (24 hpf) or 15 minutes (48 hpf), washed twice in PBST, fixed for 20 minutes in 4% PFA, and then washed five times in PBST. Following hybridization with the probe solution, the probes were saved and stored at -20°C for future use. Likewise, following the amplification stage, hairpins were saved and stored at -20°C. Recycled hairpins were heated to 95°C for 90 seconds and cooled (Bruce et al., 2021). Embryos were stored in PBS at 4°C protected from light. Whole embryos were mounted in 0.2% low melt agarose and imaged on a Leica SP8 Confocal at 10x and 40x magnification. Embryos were then genotyped following Rossi et al. (2009) with slight modifications. Following DNA extraction, PCR was performed in M buffer (2mM MgCl_2_, 13.7 mM Tris-HCl [pH 8.4], 68.4mM KCl, 0.001% gelatin, 1.8 mg/mL protease-free BSA, 136 μM each dATP/CTP/GTP/TTP) with GoTaq Flexi (Promega) and digested overnight in Fok1 enzyme at 37°C.

HCR images were first processed by performing the “rolling ball” background subtraction (50 pixels) on the sum slice projection in ImageJ (Sternberg, 1983). Signal intensity for *tbx5a* and *shha* was quantified by calculating the corrected total cell fluorescence (CTCF). Average fluorescence intensity was calculated as:

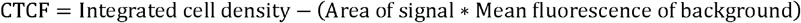

An independent, unpaired *t* test was performed to compare the CTCF and the area of expression in wildtype/heterozygotes compared to *prdm1a*^*-/-*^ mutants. Signal intensity for *fgf10a, fgf8a, dlx5a*, and *prdm1a* was quantified using the line tool in ImageJ as previously described (Fulton et al., 2020). A line was drawn from the most proximal end of the pectoral fin to the most distal tip. Signal intensity along the 30-pixel wide line was measured. The length of the fin was normalized between 0 and 1, with 0 representing the most proximal end and 1 being distal, for each individual. The signal intensity was normalized for each gene by calculating a Z-score 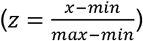. The average signal intensity along the line, the maximum intensity, and length of expression are shown. The definition of n varies for each experiment and is specified in the figure legends. Figures are 3D max projections of lateral views of the pectoral fin. Background was subtracted using the rolling ball feature in ImageJ (50 pixels).

### RNA Isolation for RT-qPCR

Total RNA was isolated from pooled wildtype/heterozygous and *prdm1a*^*-/-*^ mutant embryos after removal of the trunk at 24 and 48 hpf with TRIzol reagent (Invitrogen) and phenol/chloroform (5-10 embryo heads). RNA (500 ng) was reverse transcribed to cDNA with SuperScript III First-Strand Synthesis System (Invitrogen) for real-time quantitative PCR (RT-qPCR). Taqman primers for *prdm1a, fgf10a, fgf8a, dlx5a, dlx6a, tp63, fgf24*, and *b-actin* were purchased from Thermo Fisher Scientific. *b-actin* was used as the zebrafish internal control. Reactions were performed in at least three biological and technical replicates. Transcript abundance and relative fold change were quantified using the 2^−ΔΔCt^ method relative to control. Relative expression was compared using an unpaired, independent *t* test.

### Molecular cloning

A full-length open reading frame of *prdm1a* was amplified from cDNA as previously described (Hernandez-Lagunas et al., 2005). Amplicons were gel extracted using the Zymoclean Gel DNA Recovery Kit (Zymo Research) and recombined into the pDONR221 plasmid using BP Clonase II following the manufacturer’s instructions (Invitrogen) (Kwan et al., 2007). Sequences were confirmed with Sanger sequencing.

To delete the different protein domains of Prdm1a, unique restriction digest sites were introduced into the *pME-prdm1a* construct using the QuikChange Lightning Multi Site-Directed Mutagenesis Kit (Agilent Technologies). Primer sequences were designed using the QuikChange Primer Design Program and are included in **Table S2**. Domain deletions were modeled after *in vitro* studies described previously (Ancelin et al., 2006; Gyory et al., 2004; Su et al., 2009; Yu et al., 2000). XhoI and SalI sites were introduced to flank the SET domain; AatII sites surrounded the proline/serine rich domain; XbaI sites flanked the zinc finger domain; and XbaI sites flanked the proline/serine rich domain and zinc finger domain. Following an overnight restriction digest at 37°C with the proper enzyme, fragments were run on a 1% agarose gel, extracted, and ligated without the deleted Prdm1a domain using T4 DNA ligase (New England Biolabs) overnight at 16°C. The enzyme was inactivated at 65°C for 10 minutes before 2.5μL of the reaction was transformed into DH5α cells. Sequences were confirmed by Sanger sequencing. If sequences were out of frame, additional nucleotides were reinserted/deleted using the QuikChange Lightning kit and then resequenced (**Table S2**).

To generate *Tg(hsp70l:gal4FF)*, p5E-*hsp70l* (gift from Dr. Brian Ciruna), pME-*gal4FF* (Asakawa et al., 2008), and p3E-*polyA* were recombined into a pDestTol2CG2 destination vector using Gateway LR Clonase II (Invitrogen) (Kwan et al., 2007). The UAS constructs were created by recombining p5E-*4XnrUAS*, pME-*prdm1a* variations, and p3E-*2a-EGFP* into pDestTol2pA2 using LR Clonase II (Akitake et al., 2011; Kwan et al., 2007). Sequences were confirmed with Sanger sequencing.

### Transgenesis

Transposase mRNA was synthesized as previously described (Kawakami et al., 2004). To generate the *Tg(hsp70l:gal4FF)*^*co1025Tg*^ fish, embryos were injected at the single-cell stage with 37.5 pg of the transgene, 28.7 pg of Tol2 mRNA, and 150 mM KCl. Embryos were screened for EGFP expression in the heart at 24-72 hpf, grown to adulthood, and outcrossed to *prdm1a+/-* fish to generate stable F1 lines. Two independent *Tg(hsp70l:gal4FF); prdm1a+/-* lines were maintained. These lines were incrossed for microinjections to generate *prdm1a*^*-/-*^ mutants.

The Tol2 plasmid for generation of the *Tg(MmuPrx1:EGFP)*^*co1026Tg*^ fish that label the pectoral fin with EGFP was a generous gift from Dr. Koji Tamura (Hernandez-Vega & Minguillon, 2011; Yano & Tamura, 2013). Embryos were injected at the single-cell stage with 60 pg of transgene, 28.7 pg of Tol2 mRNA, and 150 mM KCl. Embryos were screened for EGFP expression at 24-72 hpf, grown to adulthood, and outcrossed to WT to generate stable F1 lines. Two independent *Tg(MmuPrx1:EGFP)* lines with similar expression were maintained.

### Global heat shock experiments

*Tg(hsp70l:gal4FF);prdm1a*^*+/-*^ fish were intercrossed and injected at the single-cell stage with 75 pg of the *4XnrUAS* construct, 19.1 pg of Tol2 mRNA, and 150 mM KCl. Following microinjection, embryos were heat shocked at 37°C for 60 minutes at 6 hpf. Embryos were returned to the incubator at 28.5°C to recover overnight. Embryos were then screened for mosaic EGFP expression at 24 hpf (**Fig. 6B-E)**.

### Fluorescence-activated cell sorting (FAC)

EGFP-positive pectoral fin cells were isolated from zebrafish embryos using fluorescence-activated cell (FAC) sorting. *Tg(MmuPrx1:EGFP)* embryos were collected at 24 hpf (CUT&RUN) and 48 hpf (RNA-seq) and digested in Pronase (1 mg/mL) (Roche) for 5-6 minutes to remove the chorion. Embryos were pooled together, washed in DPBS (Gibco), and dissociated in Accumax (Innovative Cell Technologies) and DNase I (50 units/100 embryos) (Roche) at 31°C for 1.5 hours. Cells were homogenized every 8 minutes by pipetting up and down using pipet tips decreasing in size. Cells were washed in solution (300 units DNase I in 4 mL DPBS) before filtering through a 70 μM nylon mesh cell strainer (Fisher Scientific) into 50 mL conical tubes pre-coated with 5% fetal bovine serum. Cells were spun down, resuspended in basic sorting buffer (1mM EDTA, 25mM HEPES [pH 7.0], 1% FBS in DPBS), stained with DAPI (1:1000), and FAC sorted on the MoFlo XDP100 sorter with a 100 μM nozzle tip.

### CUT&RUN

Following the FAC sort, cleave under targets and release using nuclease (CUT&RUN) was performed on 150,000+ sorted EGFP-positive pectoral fin cells at 24 hpf as previously described (Shull et al., 2022; Skene & Henikoff, 2017). Briefly, cells were incubated on activated Concanavalin A conjugated paramagnetic beads (EpiCypher) at room temperature for 10 minutes. Cells were washed in Antibody Buffer (20mM HEPES, pH 7.5; 150 mM NaCl; 0.5 mM Spermidine [Invitrogen], 1X Complete-Mini Protease Inhibitor tablet [Roche Diagnostics]; 0.01% Digitonin [Sigma-Aldrich]; 2 mM EDTA) and incubated overnight at 4°C with rotation in the respective antibody (IgG [1:100; Jackson ImmunoResearch, 111-005-003, RRID: AB_2337913], H3K27ac [1:66; Cell Signaling Technology, 4353S, RRID: AB10545273], and Prdm1a [1:33; rabbit polyclonal antibody from Dr. Phillip Ingham (von Hofsten et al., 2008) (Powell et al., 2013)]). Excess antibody was removed by washing in cold Digitonin Buffer (20 mM HEPES, pH 7.5; 150 mM NaCl; 0.5 mM Spermidine; 1X Complete-Mini Protease Inhibitor tablet). Cells were then incubated with pAG-MNase (EpiCypher) for 10 minutes at room temperature and washed with Digitonin Buffer. Cells were rotated in 100 mM CaCl_2_ at 4°C for two hours before the Stop Buffer (340 mM NaCl, 20 mM EDTA, 4 mM EGTA, 50 μg/ml RNaseA, 50 μg/ml glycogen) was added for 10 minutes at 37°C without the *E*.*coli* spike-in. DNA fragments were purified with a DNA Clean & Concentrate Kit (Zymo Research). Eluted DNA fragments were amplified using the NEBNext Ultra II DNA Library Prep Kit for Illumina (New England Biolabs) following the manufacturer’s instructions. Amplification of DNA was performed following guidelines outlined by EpiCypher: 98°C 45s, [98°C 15s, 60°C 10s] x14 cycles, 72°C 1 min.

Samples were subjected to paired end 150 bp sequencing on the Illumina NovaSEQ 6000 system at Novogene Corporation Inc. (Sacramento, CA). CUT&RUN experiments were performed in duplicate for two biological replicates.

### RNA-sequencing

About 150-300 *Tg(MmuPrx1:EGFP);prdm1a*^*-/-*^ (sorted by pigment phenotype (Artinger et al., 1999; Hernandez-Lagunas et al., 2005)) and *Tg(MmuPrx1:EGFP)* wildtype embryos were dissected at 48 hpf to remove the brain before FAC sorting. RNA from sorted cells was extracted using the RNAqueous™-Micro Total RNA Isolation Kit (ThermoFisher) following the manufacturer’s instructions for cultured cells. DNase treatment was performed. A library was prepared using the NEBNext Ultra II Directional RNA Library Kit for Illumina (New England Biolabs) following the manufacturer’s instructions. Samples were subjected to sequencing on the Illumina NovaSEQ 6000 system at Novogene Corporation Inc. (Sacramento, CA) at a depth of over 20 million reads per sample. RNA-seq experiments were performed in duplicate for two biological replicates per genotype.

### Bioinformatics Analysis

#### CUT&RUN

Analysis was adapted from (Ye et al., 2021). Following sequencing, paired reads were trimmed using Cutadapt (Martin, 2011). Trimmed reads were aligned to the zebrafish genome (danRer11) using Bowtie2 version 2.4.5 with the following options: --end-to-end --very-sensitive --no-mixed --no-discordant --no-unal (Langmead & Salzberg, 2012). Peak calling was performed using MACS2 2.2.7.1 using the default settings (Zhang et al., 2008), and heatmaps, bigwig tracks, and other statistics were generated with deepTools (Ramírez et al., 2014). Motif enrichment analysis was performed on peak files (bed files) using HOMER v.4.11 (Heinz et al., 2010) and the findMotifsGenome.pl script. Called peaks were annotated and subjected to GO term analysis using the ChIPseeker R package with the seq2gene function (Yu et al., 2015). Replicates were analyzed separately. There was variability between the two replicates, but they were comparable and showed similar trends.

#### RNA-seq

Following sequencing, paired reads were trimmed and mapped to the zebrafish genome (danRer11) assembly using Spliced Transcripts alignment to a Reference (STAR) v.2.7.10b (Dobin et al., 2013). Aligned counts per gene were calculated using featureCounts (Liao et al., 2014). Differential expression between wildtype and *prdm1a*^*-/-*^ was calculated using the DESeq2 package (Love et al., 2014). The top 250 differentially expressed genes by padj were plotted onto a heatmap using the pheatmaps R package (https://cran.r-project.org/web/packages/pheatmap/index.html). Gene lists were analyzed for functional annotation using GO enrichment analysis based on PANTHER Classification System (Ashburner et al., 2000; Gene Ontology, 2021; Mi et al., 2019).

## Data Availability

Sequencing data available via GEO.

https://nam02.safelinks.protection.outlook.com/?url=https%3A%2F%2Fwww.ncbi.nlm.nih.gov%2Fgeo%2Fquery%2Facc.cgi%3Facc%3DGSE217486&data=05%7C01%7Cbrittany.truong%40cuanschutz.edu%7Cb0b16ce4fac545bd05c308dac0ff6b58%7C563337caa517421aaae01aa5b414fd7f%7C0%7C0%7C638034500518463277%7CUnknown%7CTWFpbGZsb3d8eyJWIjoiMC4wLjAwMDAiLCJQIjoiV2luMzIiLCJBTiI6Ik1haWwiLCJXVCI6Mn0%3D%7C3000%7C%7C%7C&sdata=G3o50sLtqbV2ViSZHH%2FZCazdjkJ2%2FaFsn1TjWMp93nw%3D&reserved=0

## ACKNOWLEDGEMENTS

We thank members of the K.B.A laboratory for project feedback; Dr. Chris Johnson for making the *pME-prdm1a* construct; Dr. Koji Tamura for the *MmuPrx1-EGFP* plasmid; Dr. Brian Ciruna for the *p5E-hsp70l* construct; the lab of Dr. David Clouthier for cloning advice; the lab of Dr. Jamie Nichols for providing the *Tg(dlx5a:EGFP)* line; the lab of Dr. Bruce Appel for various Tol2 constructs; Dr. Lee Niswander for critically reviewing the manuscript; Christine Archer and the zebrafish fish facility team for excellent animal care; Dr. Dmitry Baturin with the University of Colorado Cancer Center Flow Cytometry Shared Resource for help with FAC sorting; Dr. Philip Ingham for the Prdm1a polyclonal antibody; Cindy Skinner for coordinating the patient studies; Dr. Elizabeth Blue and Dr. Michael Bamshad for help exome sequencing through the University of Washington Center for Mendelian Genomics (UW-CMG); and the SHFM families for participation in the study.

## COMPETING INTERESTS

We have no conflicts to declare.

## AUTHOR CONTRIBUTIONS

Conceptualization: B.T.T., C. S., H. F-S., K.B.A.; Experimentation: B.T.T., L.C.S.; Patient acquisition/identification of variants/sequencing: M.F., E.G.B., C.S., D.E., UW-CMG; Formal analysis: B.T.T., E. L.; Writing original draft: B.T.T., K.B.A.; Funding acquisition: B.T.T., C.S., H. F-S., K.B.A. Supervision: K.B.A. All authors reviewed and edited the manuscript.

## FUNDING

This work is supported by the Eunice Kennedy Shriver National Institute of Child Health and Human Development (NICHD) (1F31HD103368 to B.T.T. and 1R03HD096320-01A1 to K.B.A.). Exome sequencing by the UW-CMG was funded by the National Human Genome Research Institute (NHGRI) and the National Heart, Lung, and Blood Institute (NHLBI) (UM1 HG006493 and U24 HG008956). The content is solely the responsibility of the authors and does not necessarily represent the official views of the National Institutes of Health.

## DATA AVAILABILITY

The RNA-seq and CUT&RUN data have been deposited in NCBI’s Gene Expression Omnibus and are accessible through accession number GSE217486 (https://www.ncbi.nlm.nih.gov/geo/query/acc.cgi?acc=GSE217486).

## SUPPLEMENTARY MATERIALS

**Fig. S1: *prdm1a***^***+/-***^ **pectoral fins develop normally (related to Fig. 2)**. (**A-C**) Representative images of Alcian stained *prdm1a* (**A**) wildtype, (**B**) heterozygous, and (**C**) mutant fish at 4 dpf. There is no significant difference between wildtype and *prdm1a*^*+/-*^ in the length of the (**D**) cleithrum, (**E**) endoskeletal disk, or (**F**) fin fold or the area of the (**G**) scapulocoracoid/postcoracoid. Wildtype and heterozygote animals were combined in subsequent experiments. Measurements were averaged and compared using a one-way ANOVA, followed by a Tukey post-hoc test relative to wildtype. Error bars represent the mean ± SD. Scale bars are 50 µm. Abbreviations: cl, cleithrum; d, distal; ed, endoskeletal disk; ff, fin fold; hpf, hours post fertilization; p, proximal; pc, postcoracoid; sc, scapulocoracoid

**Fig. S2: Transient overexpression of SHFM *hPRDM1* variants in wildtype embryos suggests dominant negative effect on pectoral fin growth (related to Fig. 3)**. (**A-F**). Representative images of Alcian stained pectoral fins at 4 dpf. (**A**) Uninjected wildtype/heterozygous (n=18). (**B**) Uninjected *prdm1a*^*-/-*^ mutant (n=9). Wildtype embryos were injected with (**C**) wildtype *hPRDM1* (n=20), (**D**) *hPRDM1*(p.C239Lfs*32) (n=16), (**E**) *hPRDM1*(p.T524R) (n=14), or *hPRDM1*(p.T819A) mRNA (n=17). Measurements for the length of the (**G**) cleithrum, (**H**) endoskeletal disk, and (**I**) fin fold and (**J**) the area of the scapulocoracoid and postcoracoid were averaged and compared using a one-way ANOVA, followed by a Tukey post-hoc test relative to uninjected wildtype. Error bars represent the mean ± SD. Scale bars are 50 µm. Abbreviations: cl, cleithrum; d, distal; ed, endoskeletal disk; ff, fin fold; hpf, hours post fertilization; p, proximal; pc, postcoracoid; sc, scapulocoracoid

**Fig. S3: Overexpression of modified Prdm1a using a global heat shock Gal4/UAS system in wildtype embryos (related to Fig. 4)**. (**A-H**) Representative images of Alcian stained pectoral fins at 4 dpf are shown. (**A**) Uninjected wildtype (n=36). (**B**) Uninjected *prdm1a*^*-/-*^ mutants (n=11). Wildtype embryos were injected with *4XnrUAS-modified prdm1a-2a-EGFP* constructs containing (**C**) full-length Prdm1a (n=55), (**D**) Prdm1aΔSET (n=37), (**E**) Prdm1aΔP/S (n=38), (**F**) Prdm1aΔZnf (n=37), (**G**) Prdm1aΔP/S&Znf (n=33), and (**H**) an EGFP negative control (n=30). Scale bars are 50 µm. Measurements were taken for the length of the (**I**) cleithrum, (**J**) endoskeletal disk, and (**K**) fin fold and (**L**) the area of the scapulocoracoid and postcoracoid. Measurements for each individual were then averaged and compared using a one-way ANOVA, followed by a Tukey’s post-hoc test relative to uninjected, heat shocked *prdm1a*^*-/-*^ mutants. Error bars represent the mean ± SD. Abbreviations: Δ, deleted; cl, cleithrum; d, distal; dpf, days post fertilization; ed, endoskeletal disk; ff, fin fold; hpf, hours post fertilization; p, proximal; pc, postcoracoid; sc, scapulocoracoid

**Fig. S4: Loss of Prdm1a leads to decreased expression of limb genes (related to Fig. 6)**. (**A-L**) RT-qPCR was performed on pooled embryo heads at (**A-F**) 24 hpf and (**G-L**) 48 hpf for (**A, G**) *fgf10a*, (**B, H**) *fgf24*, (**C, I**) *tp63*, (**D, J**) *dlx5a*, (**E, K**) *dlx6a*, and (**F, L**) *fgf8a* (n=5-6 embryos per genotype per biological replicate. Three biological replicates were used). Relative expression was compared using an unpaired, independent *t* test. Error bars represent the mean ± SD. (**M-O**) Gene expression was visualized by HCR in wildtype and *prdm1a*^*-/-*^ mutants. Expression in mutants was variable. Samples with high and low expression for (**M**) *fgf10a*, (**N**) *fgf8a*, and (**O**) *shha* in mutants are shown. Scale bars are 50 µm. All images are 3D max projections of lateral views of the pectoral fin. Background was subtracted using the rolling ball feature in ImageJ (50 pixels). Abbreviations: AER, apical ectodermal ridge; d, distal; hpf, hours post fertilization; p, proximal; ZPA, zone of polarizing activity

**Fig. S5: Additional replicates from CUT&RUN showing Prdm1a directly binds to limb genes (related to Fig. 7)**. CUT&RUN was performed on isolated EGFP-positive pectoral fin cells at 24 hpf in *Tg(Mmu:Prx1-EGFP)* fish at 24 hpf. (**A-B**) Coverage heatmaps of (**A**) H3K27Ac and (**B**) Prdm1a binding across the genome 1.5 kb upstream and downstream of transcription start sight. (**C**) Annotation of enriched binding sites by Prdm1a. (**D**) Enriched Prdm1a peaks were subjected to gene ontology (GO) terms analysis using ChIPseeker’s seq2gene function. (**E**) Prdm1a peaks were subjected to motif enrichment analysis using HOMER. The top 10 motifs as well as known limb-related motifs are shown. (**F**) Tracks showing H3K27Ac enrichment (open chromatin) and Prdm1a binding sites for *fgfr1a, dlx5a/dlx6a*, and *smo*. There is variability between replicates, but the overall trends are comparable.

**Fig. S6: Hypomorphic *Tg(dlx5a:EGFP)* mutants are trending towards rescuing *prdm1a***^***-/-***^ **mutants**. (**A-F**) Representative images of Alcian stained pectoral fins at 4 dpf. Hypomorphic *Tg(dlx5a:EGFP)* fish were crossed with *prdm1a+/-* and then incrossed to assess the genetic interaction between *dlx5a* and *prdm1a*. (**A**) Wildtype (n=18). (**B**) *Tg(dlx5a:EGFP)*^*+/+*^*;prdm1a*^*-/-*^ (n=9). (**C**) *Tg(dlx5a:EGFP)*^*+/-*^*;prdm1a*^*+/+*^ (n=35). (**D**) *Tg(dlx5a:EGFP)*^*+/-*^*;prdm1a*^*-/-*^ (n=15). (**E**) *Tg(dlx5a:EGFP)*^*-/-*^*;prdm1a*^*+/+*^ (n=12) (**F**) *Tg(dlx5a:EGFP)*^*-/-*^*;prdm1a*^*-/-*^ (n=6). Measurements were taken for the length of the (**G**) cleithrum, (**H**) endoskeletal disk, and (**I**) fin fold and (**J**) the area of the scapulocoracoid and postcoracoid. Measurements for each individual were averaged and compared using a one-way ANOVA, followed by a Tukey’s post-hoc test relative to *prdm1a*^*-/-*^ mutants. Error bars represent the mean ± SD. Abbreviations: cl, cleithrum; dpf, days post fertilization; ed, endoskeletal disk; ff, fin fold; hpf, hours post fertilization; pc, postcoracoid; sc, scapulocoracoid

**Table S1: Gene candidates identified in whole exome sequencing of Split Hand/Foot Malformation patients**.

**Table S2: Primer sequences for site-directed mutagenesis**.

**Table S3: Motif enrichment analysis in called Prdm1a CUT&RUN peaks using Homer**.

## Notes

### Competing Interest Statement

The authors have declared no competing interest.

### Author Declarations

Greenwood Genetics Center, Self Regional Healthcare IRB approval for Molecular Studies of Split-Hand/Split-foot Malformations IRB#33

